# PROBABILITY OF HOSPITALIZATION AND DEATH AMONG COVID-19 PATIENTS WITH COMORBIDITY DURING OUTBREAKS OCCURRING IN MEXICO CITY

**DOI:** 10.1101/2021.12.07.21267287

**Authors:** José Sifuentes-Osornio, Ofelia Angulo-Guerrero, Guillermo De-Anda-Jáuregui, Juan L. Díaz-De-León-Santiago, Enrique Hernández-Lemus, Héctor Benítez-Pérez, Luis A. Herrera, Oliva López-Arellano, Arturo Revuelta-Herrera, Ana R. Rosales-Tapia, Rosaura Ruiz-Gutiérrez, Manuel Suárez-Lastra, Claudia Sheinbaum-Pardo, David Kershenobich

## Abstract

**Background:** Worldwide, it has been observed that there is a strong association between the severity of COVID-19 and with being over 40 years of age, having diabetes mellitus (DM), hypertension and/or obesity.

**Objective:** To compare the probability of death caused by COVID-19 in patients with comorbidities during three periods defined for this study as follows: first wave (March 23 to July 12, 2020), interwave period (July 13 to October 25, 2020), and the second wave (October 26, 2020, to March 29, 2021) using the different fatality rates observed in Mexico City.

**Methods:** The cohort studied included individuals over 20 years of age. During the first wave (symptomatic), the interwave period, and the second wave (symptomatic and asymptomatic), participants were diagnosed using nasopharyngeal swabs taken in kiosks. Symptomatic individuals with risk factors for serious disease or death were referred to hospital. SARS-CoV-2 infection was defined by real time polymerase chain reaction in all hospitalized patients. All data from hospitalized patients and outpatients were added to the SISVER database.

**Results:** The total cohort size for this study was 2,260,156 persons (having a mean age of 43.1 years). Of these, 8.6% suffered from DM, 11.6% from hypertension, and 9.7% from obesity. Of the total of 2,260,156 persons, 666,694 tested positive (29.5%) to SARS CoV-2, (with a mean age of 45). During the first wave, 82,489 tested positive; in the interwave period, 112,115; and during the second wave, 472,090. That is, a considerable increase in the number of cases of infection was observed in all age groups between the first and second waves (an increase of +472% on the first wave).

Of the infected persons, a total of 85,587 (12.8%) were hospitalized: 24,023 in the first wave (29.1% of those who tested positive in this period); 16,935 (15.1%) during the interwave period, and 44,629 (9.5%) in the second wave, which represents an increase of 85.77% on the first wave.

Of the hospitalized patients, there were 42,979 deaths (50.2% of those hospitalized), in the first wave, 11,964 (49.8% of those hospitalized in this period), during the interwave period, 6,794 (40.1%), and in the second wave 24,221 (54.3%), an increase of +102.4% between the first wave and the second.

While within the general population, the probability of a patient dying having both COVID-19 and one of the specified comorbidities (DM, obesity, or arterial hypertension) showed a systematic reduction across all age groups, the probability of death for a hospitalized patient with comorbidities increased across all age groups during the second wave. When comparing the fatality rate of hospitalized COVID-19 patients in the second wave with those of the first wave and the interwave period, a significant increase was observed across all age groups, even in individuals without comorbidities.

**Conclusion:** The data from this study show a considerable increase in the number of detected cases of infection in all age groups between the first and second waves. In addition, 12.8% of those infected were hospitalized for severe COVID-19, representing an increase of +85.9% from the first wave to the second. A high mortality rate was observed among hospitalized patients (>50%), as was a higher probability of death in hospitalized COVID-19 patients with comorbidities for all age groups during the second wave, although there had been a slight decrease during the interwave period.

**SUMMARY BOX:** *What is already known?:* Worldwide the resurging of COVID-19 cases in waves has been observed. In Mexico, like in the rest of the world, we have observed surges of SARS CoV-2 infections, COVID-19 hospitalizations and fatal outcomes followed by decreases leading to local minima. Pre-existing health conditions such as being older, having diabetes mellitus (DM), hypertension and/or obesity has been observed to be associated with an increase in the severity of COVID-19.

*What are the new findings?:*

1. Between the first and second waves, considerable increases were observed in the number of detected cases of infection (+472%), in the number of hospitalized subjects (+85.9%), and the number of hospitalized subjects and deaths (+102.4%) in all age groups.
2. When analysing only hospitalized individuals, with or without comorbidities, the Case Fatality Rate was high (50.2%), the probability of death increased considerably in all age groups between the first and second waves. This increase was more noticeable in those individuals with previously identified comorbidities (DM, hypertension, or obesity).
3. An increased probability of death among individuals without comorbidities was observed between the first and second waves.

*What do the new findings imply?:* During the second wave, demand for hospitalization increased, magnifying the impact of age and comorbidities as risk factors. This situation highlights the importance of decreasing the prevalence of comorbidities among the population.

## INTRODUCTION

The original outbreak of SARS-CoV-2 occurred in the province of Wuhan, China, in November 2019.[1] It then spread rapidly worldwide and was categorized as a pandemic by the WHO in 2020.[2] During the progress of the pandemic in several regions of the world, it was observed that being elderly, having chronic obstructive pulmonary disease, diabetes mellitus (DM), arterial hypertension, or obesity were common features among individuals who required hospitalization or died and were associated with increased risk for the development of the disease [3] caused by the SARS-CoV-2 virus (COVID-19).

This association has been systematically observed throughout the world, the basis of which seems to be the pro-inflammatory state suffered by patients living with these chronic non-communicable diseases.[4] It has also been observed that patients with uncontrolled hypertension develop an excess of receptors ACE-2.[5] Since these conditions are common in adults around the world, obesity and being overweight particularly, as well as uncontrolled or undiagnosed DM or hypertension, COVID-19 has found fertile ground in which to take root and spread. The disease has a good chance of permanently residing in this environment and infecting other individuals with poor or limited immunity mechanisms, such as people with organ transplants, patients with neoplastic diseases undergoing chemotherapy, patients with chronic rheumatic diseases under immunosuppressive treatment, among other groups.[6]

The appearance of COVID-19 in the previously healthy population has attracted much attention recently since this disease has been observed in children, adolescents and young adults who are at risk of death or serious deterioration, in addition to the risks of kidney, lung, or liver complications, which can lead to permanent disability. It has also been observed that the risk of complications increases with the number of pre-existing comorbidities. Complications tend to be more frequent in men than in women, and in those patients who were admitted to intensive care units or required mechanical ventilation.[7] The pre-existence of overweight/obesity, DM, or hypertension (8) has meant that these complications have been observed frequently in the Mexican population. The prevalence of DM has been reported in adults over 20 years of age at rates of 10.3% throughout the country and 12.7% in Mexico City (CDMX); in the case of hypertension, 18.4% and 20.2%, respectively. While in the case of obesity, the figures are 75.2% (39.1% overweight and 36.1% obesity) and 75.9% (overweight 40.6% and obesity 35.3%), for the country as a whole and for Mexico City, respectively.[8]

The Mexico City Government has implemented various precautionary measures in the population based on the recommendations of the World Health Organization, the Federal Government, and the Metropolitan Committee on Health, [9-11] see Supplementary Tables 1, 2, and 3. In summary, these were:

1. Application of an epidemiological mobility/risk of transmission indicator, the Epidemiological ‘Traffic Light’, coded green, yellow, orange, and red for carrying out school, commercial, artistic, sports activities, etc. in CDMX. Code green indicates normal mobility with 0% risk of transmission and code red indicates the maximum application of containment measures corresponding to a >90% risk of transmission. The distribution of the different colours (risks) during the study period can be seen in Table S1.
2. Suspension of classes, activities in shopping centres, shows, restaurants and meeting places, places of entertainment or worship, and all kinds of face-to-face activities in the offices and dependencies of the CDMX government.
3. Promotion of the use of face masks while in public places and frequent handwashing with soap and water or at minimum with gel alcohol.
4. Provision of a medical guidance service for people presenting with symptoms of respiratory illness (fever, cough, sore throat, headache, tiredness), including the instruction to stay at home and send an SMS text message saying ‘COVID-19’ to the number 51515, support via the LOCATEL information service (55-5658-1111), on the CDMX website and via Facebook and via periodic monitoring.
5. Detection and follow-up of patient contacts.
6. Opening of several acute care hospitals for patients with COVID-19 and hiring of medical personnel.
7. Ambulance service to transfer patients from home to hospitals and referral between hospitals.
8. Opening Announcement of the first stage of the Temporary Unit to care for patients with COVID-19 at the Centro CitiBanamex supported by private donations.
9. Oxygen program in your home in collaboration with a private company (INFRA S.A. de C.V.), see Supplementary Tables S2 and S3.

In prior studies carried out by other Mexican groups [12,13] and in a study carried out by this group in the CDMX, [Sifuentes-Osornio J, Angulo-Guerrero O, Benítez-Pérez H, et al. Unpublished observations], a strong association between the severity of COVID-19, being over 40 years of age, and certain non-communicable chronic diseases, in particular DM, high blood pressure and obesity, was observed. This group has previously analysed the impact of comorbidities and different age ranges on the risk of developing complications. However, the evolution of the pandemic suggests the possibility that the changes could have occurred as a result of the interventions, as well as being due to changes in population dynamics, for example, mobility or the emergence of variants of concern in the country, among other conditions. [14,15] Therefore, the objective of this study was to compare the probability of death caused by COVID-19 in patients with comorbidities during three periods of the pandemic that we defined as first-wave (March 23 to July 12, 2020), interwave period (July 13 to October 25, 2020), and second-wave (October 26, 2020, to March 29, 2021), based on the different Case Fatality Rates (CFR) that we observed in the CDMX during the study period.

## MATERIALS AND METHODS

### Study population and design

This retrospective observational study included individuals aged over 20: 193,579 people tested during the first wave; 338,639 during the interwave period, and 1,727,938 during the second wave. All the subjects included were evaluated at the outpatient clinics, emergency services, strategically located health kiosks, and hospitalization centres of the health institutions of the CDMX between March 23, 2020, and March 29, 2021. All the people included during the first wave presented with one or more of the following symptoms: fever, cough, sore throat, dyspnoea, rhinorrhoea, nasal congestion, conjunctivitis, myalgia, arthralgia, headache, for more than one day. The people included during the interwave period, and the second wave included both symptomatic and asymptomatic individuals whose samples were taken in specially established kiosks in different locations in CDMX. The study was approved by the Research and Research Ethics Committees of the Instituto Nacional de Ciencias Médicas y Nutrición Salvador Zubirán (IRB: 3347).

In each case, information was obtained on demographic data, recent trips (during the first wave), contact with other suspected or confirmed cases, comorbidities, signs, and symptoms, and monitoring of the evolution of their illness, using a COVID-19 epidemiological case study format prepared by the Ministry of Health (supplementary material). Data on the dates of hospitalization and death were included as an outcome. Individuals who tested positive on the diagnostic test and having one or more risk factors for serious disease or death (age ≥65 years, systemic arterial hypertension, DM mellitus, obesity, chronic obstructive pulmonary diseases (COPD), ischemic heart disease, cardiovascular disease (CVD), temperature ≥39°C, respiratory rate ≥24 rpm, saturation O2 <92%, dyspnoea, abdominal pain) were referred to the reference institutions participating in the program for a thorough evaluation and better care.

### Definition of waves and interwave period

In Mexico, the first identified case of COVID-19 occurred on February 27, 2020, and by March 18, the first death from that cause was recorded. Five days later, on March 23, 2020, Mexico announced the start of the quarantine, and the decision was made to close schools and suspend nonessential activities. A week later, on April 2, 2020, a state of emergency was declared throughout the country. During this initial phase of the pandemic, Mexico ranked third in the world in the number of infections and deaths recorded, behind the USA and Brazil.

To avoid using a biased definition of the study periods, we defined a wave as the period between two local minima in the time series of epidemiologic parameters (that is, the second derivative of the time series is zero). To avoid biases from changes in testing protocols, we used the time series of weekly new confirmed hospitalized cases and weekly deaths in hospitals. We then defined the interwave as the period between the end of a wave and the start of another.

### Diagnostic procedures

SARS-CoV-2 infection was detected by real-time reverse-polymerase chain reaction (RT-qPCR) with reverse transcriptase by Applied Biosystems 7500 (Applied Biosystems, Foster City, CA, USA), exclusively, between March 23 and November 30, 2020, as previously described. [16] Subsequently, the SARS-CoV-2 Roche SARS-CoV-2 rapid antigen test (Roche, Basel, Switzerland) or Abbott BinaxNOWTM 88 COVID-19 Ag Card (Abbott Laboratories, Abbott Park, IL, USA) determination was used for antigen detection in the nasopharyngeal swab following the manufacturer’s recommendations, between December 1, 2020, and March 29, 2021. In all hospitalized patients, the diagnosis of COVID-19 was confirmed by RT-qPCR.

All data from hospitalized patients and outpatients residing in the CDMX were added to the Epidemiological Surveillance System for Respiratory Diseases (SISVER) database [17] for each medical unit and included demographic data, medical unit, results of the SARS CoV-2 diagnostic test, residence, age, sex, date of onset of symptoms, date of admission to hospital, comorbidities, and date of death, among others. Additionally, the CDMX Government’s Digital Agency for Public Innovation completed the data, incorporating detailed information on the cases, as well as other indicators such as the number of beds available for COVID-19 care, hospitalization rates, and overall availability of beds.

### Statistical analysis

Considering that, in the context of COVID-19, the presence of certain comorbidities has been associated with more unfavourable outcomes, such as hospitalization, intubation, and death, a classification model was implemented that identifies individual patient’s risk to either outcome, based on their associated comorbidities. For that, the use of conditional probabilities over descriptive statistics was chosen, since the former are usually less susceptible to sampling errors and normalize the results to the conditions given by the independent variables of interest. This allows us to answer questions that are of interest to the individual. So, if the fact of interest is the likelihood of death when presenting with DM and suffering from COVID-19, there is a conditional probability for obtaining the answer to that specific question for both the event of interest (dependent variable) and for the chosen precondition (independent variable). Then, the conditional probability of event A given event B is obtained as follows: *P(A\B) = P(A⍰B)/P(B)*, and when we are interested in multiple events or conditions, the conditional probability is obtained as follows: *P(A\B1⋀B2⋀*… *⋀Bn)=P(A⍰B1B2⍰*… *⍰Bn)/P(B1B2⍰*… *⍰Bn)*.[18]

In this study, an analysis for each 20-year age bracket was generated for each period: first wave, the interwave period, and the second wave. In each age bracket, a Bayesian analysis was performed on the three comorbidities: DM, hypertension, and obesity. [19] Although the SISVER database may have some ‘measurement uncertainty’, the Bayesian model is not affected by this kind of ‘uncertainty’ since the potential biases are incorporated into the priors from the start. Thus, the resulting analysis model was ‘strengthened’ by using this strategy.

For all cases, the following conditional probabilities were calculated: a) the probability of death given that the person was positive for SARS-CoV-2 infection and had the comorbidity (supplementary material), and b) the probability of death given that the person developed COVID-19, was hospitalized, and had the comorbidity. Furthermore, the same conditional probabilities were obtained for patients without any of the following comorbidities: DM, hypertension, obesity, COPD, cardiac diseases, or renal failure. These patients were considered as the healthy population baseline for this study. Finally, the conditional probabilities for each time frame were compared to identify what had changed in time for every case. It is important to point out that all the conditional probabilities share a base condition, namely the SARS-CoV-2 testing that was performed on many individuals who voluntarily attended the kiosks due to the recent appearance of symptoms or their own suspicion of having an asymptomatic infection.

Regarding the sensitivity of the Bayesian analysis, the precision in the results is inversely proportional to the frequency of the conditional event. For instance, if the conditional event B has a frequency of 1000, it can be expected to have a maximum uncertainty of 0.1% peak to peak, or a mean uncertainty of plus or minus 0.0005, in all the probabilities conditioned to B.[19]

### Comparative analysis of lethality in hospitalized patients during the waves in contrast to the interwave period

We used Fisher’s Exact Test to evaluate whether the proportion of survivors and deaths among hospitalized patients showed differences between the periods evaluated. Since the ratio between survivors and deaths is proportional to the CFR, we concluded that the CFR among hospitalized patients changed during the periods of greater transmission and greater demand for hospitalization.

The following contrasts were made: 1) Comparison between hospitalized patients during either of the two waves and in the interwave period and 2) Comparison between hospitalized patients in the first peak and the second peak. The comparison was made using Fisher’s exact test, considering the following contingency table:

|  | Death | No death |
| --- | --- | --- |
| Period A | The death count in Period A | Survivor (cases that did not end with death) count in Period A |
| Period B | The death count in Period B | Survivor (cases that did not end with death) count in Period B |

The p-values of the multiple comparisons were corrected using the False Discovery Rate (FDR), and an FDR value <0.05 was considered significant.

To avoid Simpson Paradox issues that might obscure differences, these comparisons were performed in subpopulations defined by age group and the combinations of the three comorbidities studied; additionally, the subpopulation of patients not presenting with any of the three specified comorbidities but with any other recorded comorbidity was studied separately from the subpopulation with no recorded comorbidity. We additionally, compared the interwave period with the two waves to gain a better notion of changes in lethality associated with periods of increased transmission and associated with greater demand for hospitalization. Thus, one of our objectives was to evaluate whether there were changes in lethality between the interwave period and the second wave. The rationale was that while in the first wave knowledge of the disease was scarce and detection programmes were being developed, by the interwave period, several interventions had been adopted, including increased testing programmes. Given that lethality increases during periods of higher transmission and demand for hospitalization, we calculated the Pearson correlation between weekly CFR for hospitalized patients and hospital occupancy as reported. [20]

### Correlation between weekly hospitalized Case Fatality Rate and hospital capacity

We calculated the Pearson correlation between weekly Case Fatality Rates (CFR) for hospitalized patients in each age group *versus* hospital capacity (as recorded in the federal dashboard). [20]

## RESULTS

### Descriptive analysis

During the study period, a cohort of 2,260,156 subjects were included in the study, all having been tested for SARS-CoV-2 infection performed by nasopharyngeal RTq-PCR or antigen testing. The mean age of the subjects was 43.1 years (SD 15.0), 1,172,907 (51.9%) being women. The cohort included 194,647 individuals suffering from DM (8.6%); 262,652 had hypertension (11.6%), and 218,769 were obese (9.7%). Of the total of studied subjects, 666,694 tested positive (29.5%) to SARS CoV-2, having a mean age of 45.0 (SD 15.6). Of those testing positive, 340,811 (51.1%) were women. There was a considerable increase in the number of cases (+472%) of infection in all age groups between the first wave and the second. See Table 1. The number of subjects in each age bracket was as follows: 20 to 39 years of age: 273,715 (41% of whom tested positive), 140,248 being women (51.1%); 40 to 59 years of age: 269,285 (40.4% of whom tested positive), 139,241 being women (51.7%); 60 to 79 years of age: 109,467 (16.4% of whom tested positive) 53,964 being women (49.3%); 80 years of age and over: 14,227 (2.1% of whom tested positive), 7,358 being women (51.7%).

**Table 1:**
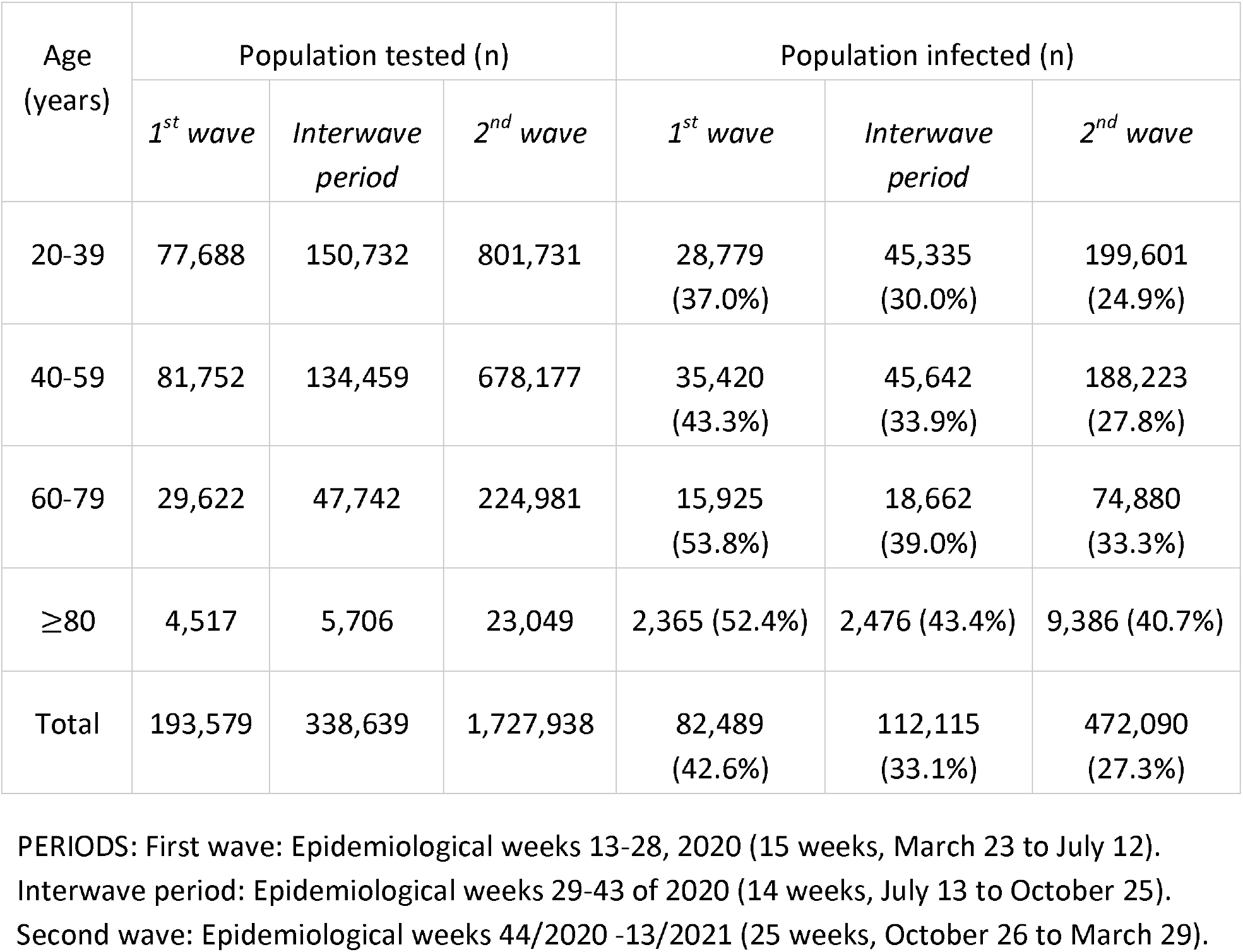
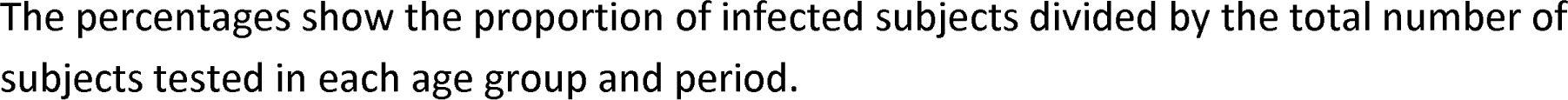
Sample size of the population tested and infected by SARS-CoV-2 by age groups and COVID-19 waves.

| Age (years) | Population tested (n) |  |  | Population infected (n) |  |  |
| --- | --- | --- | --- | --- | --- | --- |
|  | *1<sup>st</sup> wave* | *Interwave period* | *2<sup>nd</sup> wave* | *1<sup>st</sup> wave* | *Interwave period* | *2<sup>nd</sup> wave* |
| 20-39 | 77,688 | 150,732 | 801,731 | 28,779<br>(37.0%) | 45,335<br>(30.0%) | 199,601<br>(24.9%) |
| 40-59 | 81,752 | 134,459 | 678,177 | 35,420<br>(43.3%) | 45,642<br>(33.9%) | 188,223<br>(27.8%) |
| 60-79 | 29,622 | 47,742 | 224,981 | 15,925<br>(53.8%) | 18,662<br>(39.0%) | 74,880<br>(33.3%) |
| ≥80 | 4,517 | 5,706 | 23,049 | 2,365 (52.4%) | 2,476 (43.4%) | 9,386 (40.7%) |
| Total | 193,579 | 338,639 | 1,727,938 | 82,489<br>(42.6%) | 112,115<br>(33.1%) | 472,090<br>(27.3%) |

PERIODS: First wave: Epidemiological weeks 13-28, 2020 (15 weeks, March 23 to July 12).
Interwave period: Epidemiological weeks 29-43 of 2020 (14 weeks, July 13 to October 25).
Second wave: Epidemiological weeks 44/2020 -13/2021 (25 weeks, October 26 to March 29).
The percentages show the proportion of infected subjects divided by the total number of subjects tested in each age group and period.

Of the total of infected individuals, 85,587 (12.8%) were hospitalized after being diagnosed as having severe respiratory acute infection. This total breaks down as follows: 24,023 of 82,489 infected people (29.1%) during the first wave; 16,935 of 112,115 (15.1%) during the interwave period, and 44,629 of 472,090 (9.5%) during the second wave.

Of the hospitalized subjects, there was an increase of +85.9% between waves, of +36.2% in the case of the group aged 20 to 39 years, +58.6% for the group aged 40 to 59, +121.2% for the group aged 60 to 79 years, and +171.4% for the 80 years or older group, see Table 2. This shows that the relative increase in hospitalizations between the first wave and the second increased linearly as a function of age groups, as seen in Figure 1.

**Table 2:**
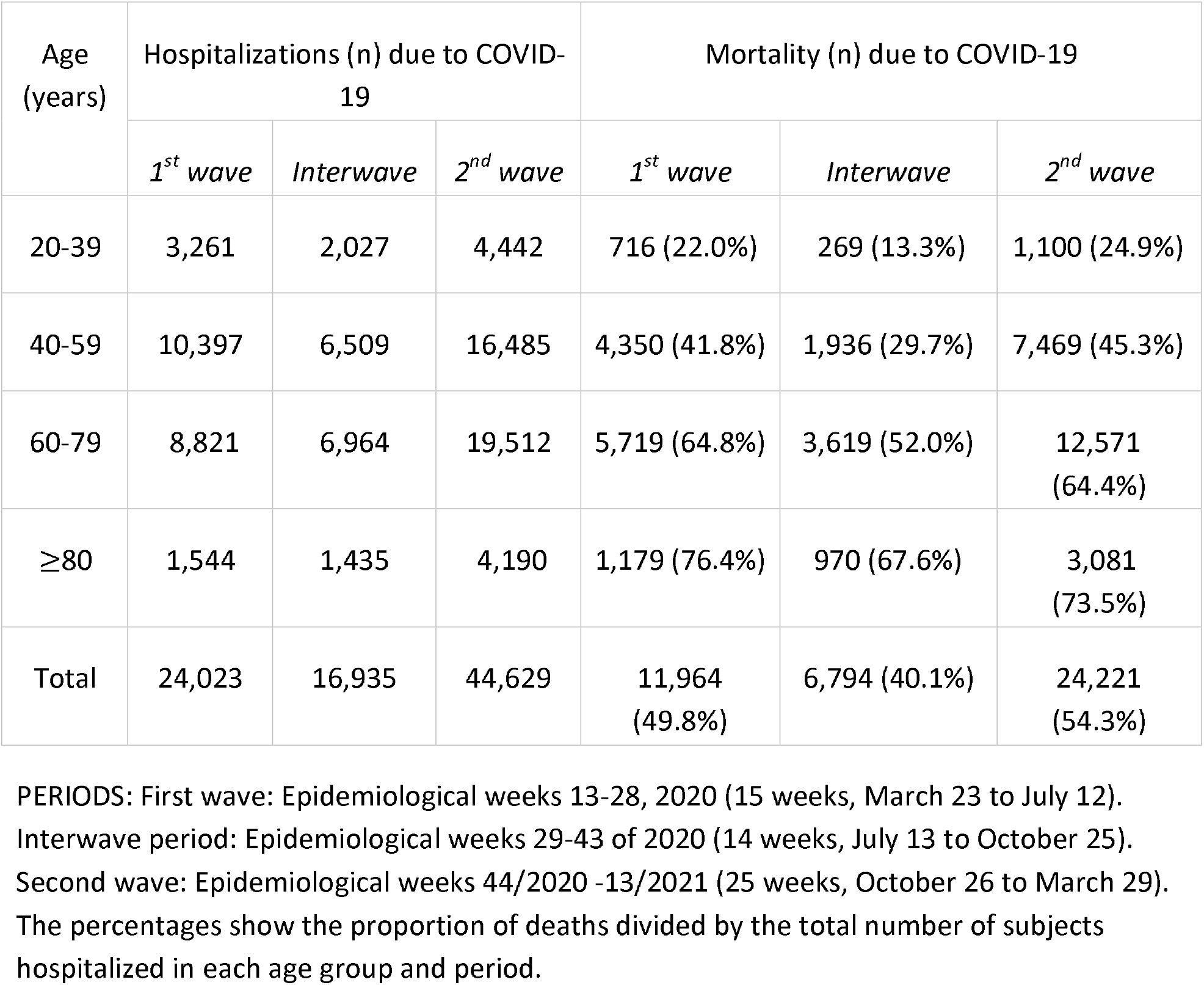
Sample size of hospitalized patients and mortality by COVID-19 by age groups and pandemic waves.

**Figure 1.**
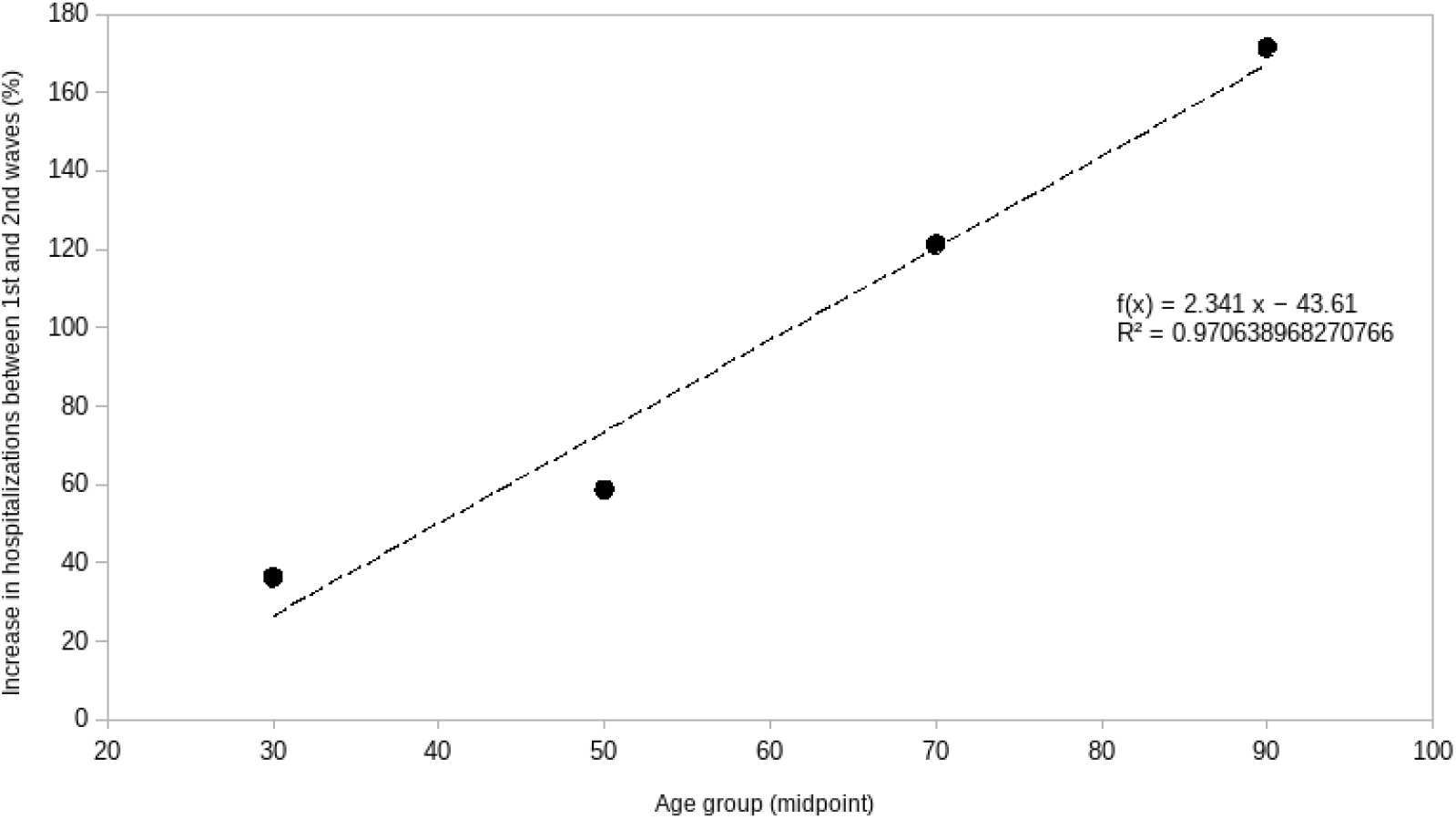
An increase in the number of hospitalizations was observed with age group brackets

### Comparative analysis of the lethality in hospitalized patients during the waves in contrast with the interwave period

During the study, there were 42,979 deaths among the 85,587 hospitalized individuals (50.2%), being 11,964 in the first wave, 6,794 in the interwave period, and 24,221 in the second wave, an increase of +102.4% between the two waves. By age, the percentage increases were: +53.6% in the case of the group aged 20 to 39, +71.7% for the group aged 40 to 59, +119.8% for the group aged 60 to 79, and +161.3% for the group aged 80 and older, see Table 2. Given that both the numbers of tests performed, and subjects tested were notably greater during the interwave period and during the second wave (Table 1), the probability of death was significantly smaller in all age groups of the general population, as shown in Table 3. In contrast, the probability of death among individuals hospitalized for COVID-19 with or without comorbidities increased systematically in all age groups, except for the group aged 60 to 79, which remained constantly high, see Table 4.

**Table 3:**
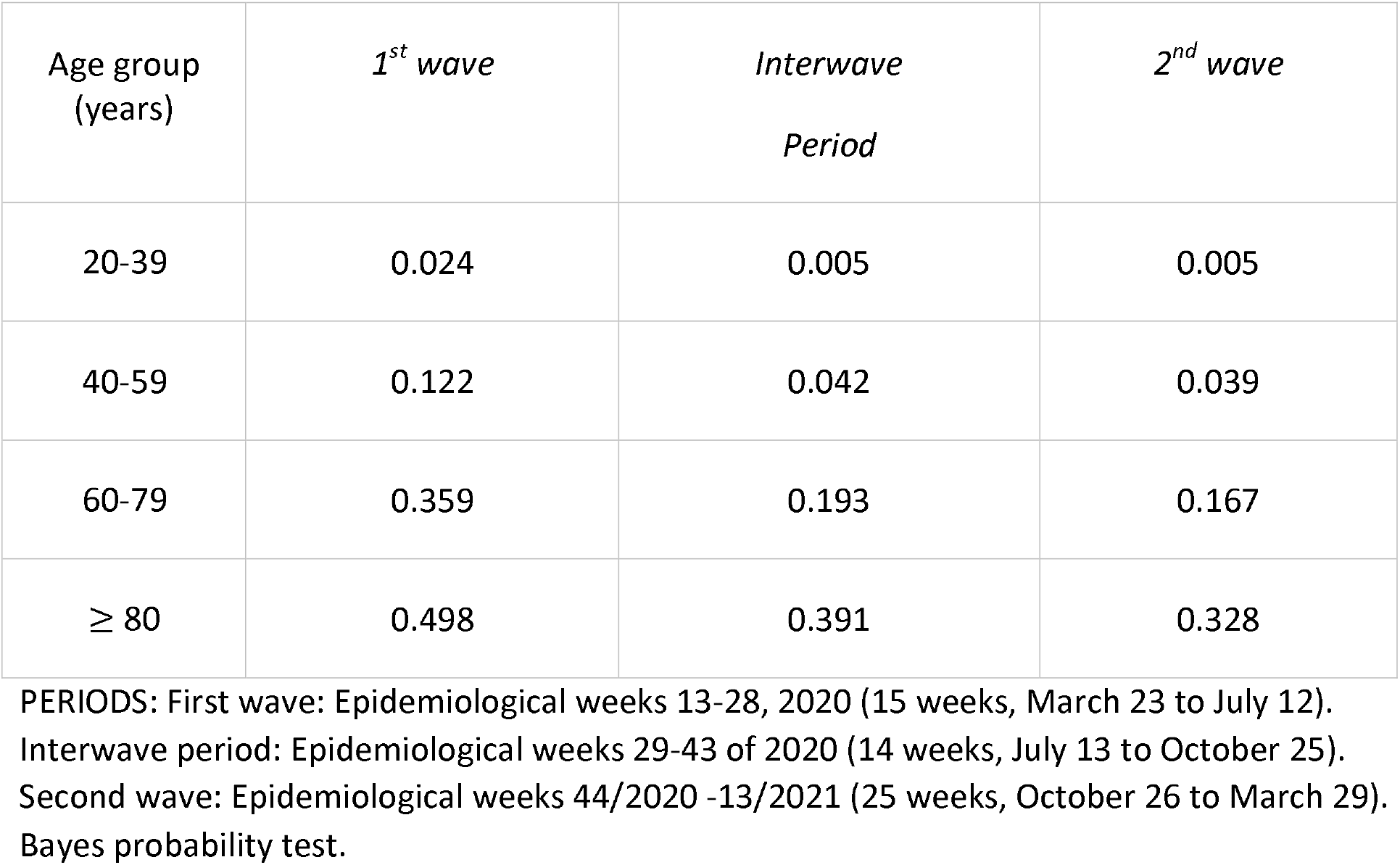
Global lethality probability of COVID-19 patients by age group and period.

| Age group<br>(years) | *1<sup>st</sup> wave* | *Interwave*<br>*Period* | *2<sup>nd</sup> wave* |
| --- | --- | --- | --- |
| 20-39 | 0.024 | 0.005 | 0.005 |
| 40-59 | 0.122 | 0.042 | 0.039 |
| 60-79 | 0.359 | 0.193 | 0.167 |
| ≥ 80 | 0.498 | 0.391 | 0.328 |

PERIODS: First wave: Epidemiological weeks 13-28, 2020 (15 weeks, March 23 to July 12). Interwave period: Epidemiological weeks 29-43 of 2020 (14 weeks, July 13 to October 25). Second wave: Epidemiological weeks 44/2020 -13/2021 (25 weeks, October 26 to March 29). Bayes probability test.

**Table 4:** Mortality probability of hospitalized COVID-19 patients by age group and period.

| Age group (years) | *1<sup>st</sup> wave* | *Interwave period* | *2<sup>nd</sup> wave* |
| --- | --- | --- | --- |
| 20-39 | 0.185 | 0.116 | 0.228 |
| 40-59 | 0.368 | 0.274 | 0.418 |
| 60-79 | 0.577 | 0.486 | 0.560 |
| ≥80 | 0.673 | 0.630 | 0.682 |

PERIODS: First wave: Epidemiological weeks 13-28, 2020 (15 weeks, March 23 to July 12).
Interwave period: Epidemiological weeks 29-43 of 2020 (14 weeks, July 13 to October 25).
Second wave: Epidemiological weeks 44/2020 -13/2021 (25 weeks, October 26 to March 29).
Bayes probability test

The probability of dying among individuals suffering from DM mellitus, obesity, or arterial hypertension and COVID-19 in the general population showed a systematic reduction in all age groups, see supplementary Tables S5, S6, S7, S8, S9, S10, S11, S12, S13, S14, S15, S16. In contrast, the probability of dying for hospitalized individuals with DM (Table 5), who suffered from obesity (Table 6), or who suffered from hypertension (Table 7) increased in all age groups systematically during the second wave.

**Table 5:** Mortality probability of hospitalized COVID-19 patients with diabetes mellitus by age group and period.

| Age group (years) | *1<sup>st</sup> wave* | *Interwave period* | *2<sup>nd</sup> wave* |
| --- | --- | --- | --- |
| 20-39 | 0.338 | 0.210 | 0.366 |
| 40-59 | 0.452 | 0.347 | 0.481 |
| 60-79 | 0.596 | 0.524 | 0.619 |
| ≥80 | 0.678 | 0.640 | 0.684 |

PERIODS: First wave: Epidemiological weeks 13-28, 2020 (15 weeks, March 23 to July 12).
Interwave period: Epidemiological weeks 29-43 of 2020 (14 weeks, July 13 to October 25).
Second wave: Epidemiological weeks 44/2020 -13/2021 (25 weeks, October 26 to March 29).
Bayes probability test.

**Table 6:** Mortality probability of hospitalized COVID-19 patients with obesity by age group and period.

| Age group (years) | *1<sup>st</sup> wave* | *Interwave period* | *2<sup>nd</sup> wave* |
| --- | --- | --- | --- |
| 20-39 | 0.252 | 0.161 | 0.272 |
| 40-59 | 0.387 | 0.264 | 0.462 |
| 60-79 | 0.572 | 0.489 | 0.617 |
| ≥80 | 0.684 | 0.667 | 0.688 |

PERIODS: First wave: Epidemiological weeks 13-28, 2020 (15 weeks, March 23 to July 12).
Interwave period: Epidemiological weeks 29-43 of 2020 (14 weeks, July 13 to October 25).
Second wave: Epidemiological weeks 44/2020 -13/2021 (25 weeks, October 26 to March 29).
Bayes probability test.

**Table 7:** Mortality probability of hospitalized COVID-19 patients with hypertension by age group and period.

| Age group (years) | 1<sup>st</sup> wave | Interwave period | 2<sup>nd</sup> wave |
| --- | --- | --- | --- |
| 20-39 | 0.316 | 0.227 | 0.402 |
| 40-59 | 0.428 | 0.334 | 0.483 |
| 60-79 | 0.586 | 0.513 | 0.611 |
| ≥80 | 0.674 | 0.660 | 0.687 |

PERIODS: First wave: Epidemiological weeks 13-28, 2020 (15 weeks, March 23 to July 12).
Interwave period: Epidemiological weeks 29-43 of 2020 (14 weeks, July 13 to October 25).
Second wave: Epidemiological weeks 44/2020 -13/2021 (25 weeks, October 26 to March 29).
Bayes probability test.

### Differences in lethality among hospitalized patients with COVID-19 during waves and in the interwave period

We used the Fisher Exact Test to identify whether individual hospitalized populations exhibited changes in lethality as the pandemic evolved. The first comparison contrasted lethality in the interwave period with that of the two waves. Table 8 shows changes in Case Fatality Rates (CFR) for hospitalized patients with the subpopulations found significant using Fisher’s exact test (FDR < 0.05). We observed increases in lethality across all age groups. In each age group, the subpopulation ranked first, in terms of FDR, was the one composed of patients without comorbidities. However, in terms of the magnitude of the lethality increase (in terms of the difference in CFR), each age group behaved differently. In the case of the 20 to 39 group, the largest increase was found in the subpopulation with DM and hypertension; in the 40 to 59 group, the subpopulations with DM, hypertension, and obesity showed the largest increase; in the 60 to 79 group the subpopulation with obesity registered the highest increase; finally, in patients aged 80 and over, the largest increase in lethality was in the subpopulation without recorded comorbidities. When comparing lethality between the first and second waves, only the subpopulation of patients aged 40 to 59 without comorbidities showed a significant (FDR = 4.46E-02) yet small (change in CFR = 2.09) increase.

**Table 8.** Lethality comparison between waves and the interwave period among hospitalized patients with COVID-19.

| Age group | Comorbidities | CFR-interwave | CFR-waves | Difference-CFR | Fisher exact test FDR |
| --- | --- | --- | --- | --- | --- |
| [20,39] | none | 7.91 | 11.65 | 3.74 | 1.42E-05 |
| [20,39] | obes | 13.00 | 18.21 | 5.21 | 1.50E-02 |
| [20,39] | dm; hypert | 16.07 | 32.55 | 16.48 | 3.37E-02 |
| [20,39] | dm; obes | 13.46 | 28.76 | 15.30 | 4.48E-02 |
| [40,59] | none | 20.37 | 29.72 | 9.34 | 2.29E-27 |
| [40,59] | obes | 22.35 | 33.79 | 11.45 | 1.30E-11 |
| [40,59) | dm; hypert;<br>obes | 27.72 | 42.02 | 14.30 | 1.65E-06 |
| [40,59) | hypert | 25.57 | 35.33 | 9.76 | 2.35E-06 |
| [40,59) | dm; hypert | 32.86 | 42.33 | 9.47 | 2.35E-06 |
| [40,59) | hypert; obes | 26.04 | 37.69 | 11.64 | 5.00E-05 |
| [40,59) | dm | 30.10 | 38.08 | 7.98 | 6.81E-05 |
| [40,59) | dm; obes | 26.72 | 38.52 | 11.80 | 1.20E-03 |
| [40,59) | other | 24.36 | 29.15 | 4.79 | 2.51E-02 |
| [60,79) | none | 40.85 | 49.89 | 9.04 | 8.82E-16 |
| [60,79) | dm | 41.51 | 53.41 | 11.90 | 3.35E-10 |
| [60,79) | hypert | 41.01 | 50.55 | 9.54 | 3.64E-09 |
| [60,79) | obes | 39.41 | 52.80 | 13.39 | 2.35E-06 |
| [60,79) | dm; hypert | 46.79 | 53.64 | 6.84 | 2.35E-06 |
| [60,79) | other | 39.07 | 47.82 | 8.75 | 1.02E-04 |
| [60,79) | dm; hypert;<br>obes | 45.36 | 54.21 | 8.85 | 4.10E-04 |
| [60,79) | dm; obes | 39.81 | 54.26 | 14.45 | 7.39E-04 |
| [60,79) | hypert; obes | 46.10 | 52.27 | 6.16 | 4.48E-02 |
| [80+) | None | 50.47 | 59.02 | 8.55 | 8.76E-04 |
| [80+) | dm; hypert | 51.75 | 58.02 | 6.27 | 4.48E-02 |

dm: diabetes mellitus; hypert: arterial hypertension; obes: obesity; CFR-wave: cases occurred in either peak; difference-CFR: the difference between CFR-wave and CFR-interwave; CFR-interwave: cases occurred during the period interpeak; statistically significant value: FDR<0.05. Periods: First wave: Epidemiological weeks 13-28, 2020 (15 weeks, March 23 to July 12). Interwave period: Epidemiological weeks 29-43 of 2020 (14 weeks, July 13 to October 25). Second wave: Epidemiological weeks 44/2020 -13/2021 (25 weeks, October 26 to March 29). Fisher exact test with BH correction used to estimate significance.

Finally, comparing the interwave period with the waves gave us a notion of changes in lethality associated with periods of increased transmission and demand for hospitalization. Table 9 shows the results of this analysis. We observed statistically significant increases in several subpopulations (23 out of 48 evaluated), which again highlighted higher lethality in the period with increased hospital demand. Nevertheless, it should be noted that when comparing the global population of hospitalized and ambulatory COVID-19 patients, we do see some populations with significant decreases in lethality (see Supplementary Table 4), particularly in persons older than 60 years of age and younger populations with an increased risk of lethality.

**Table 9.**
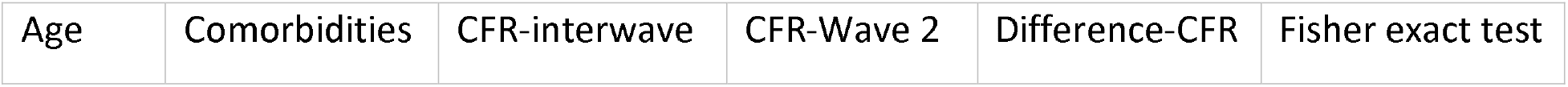
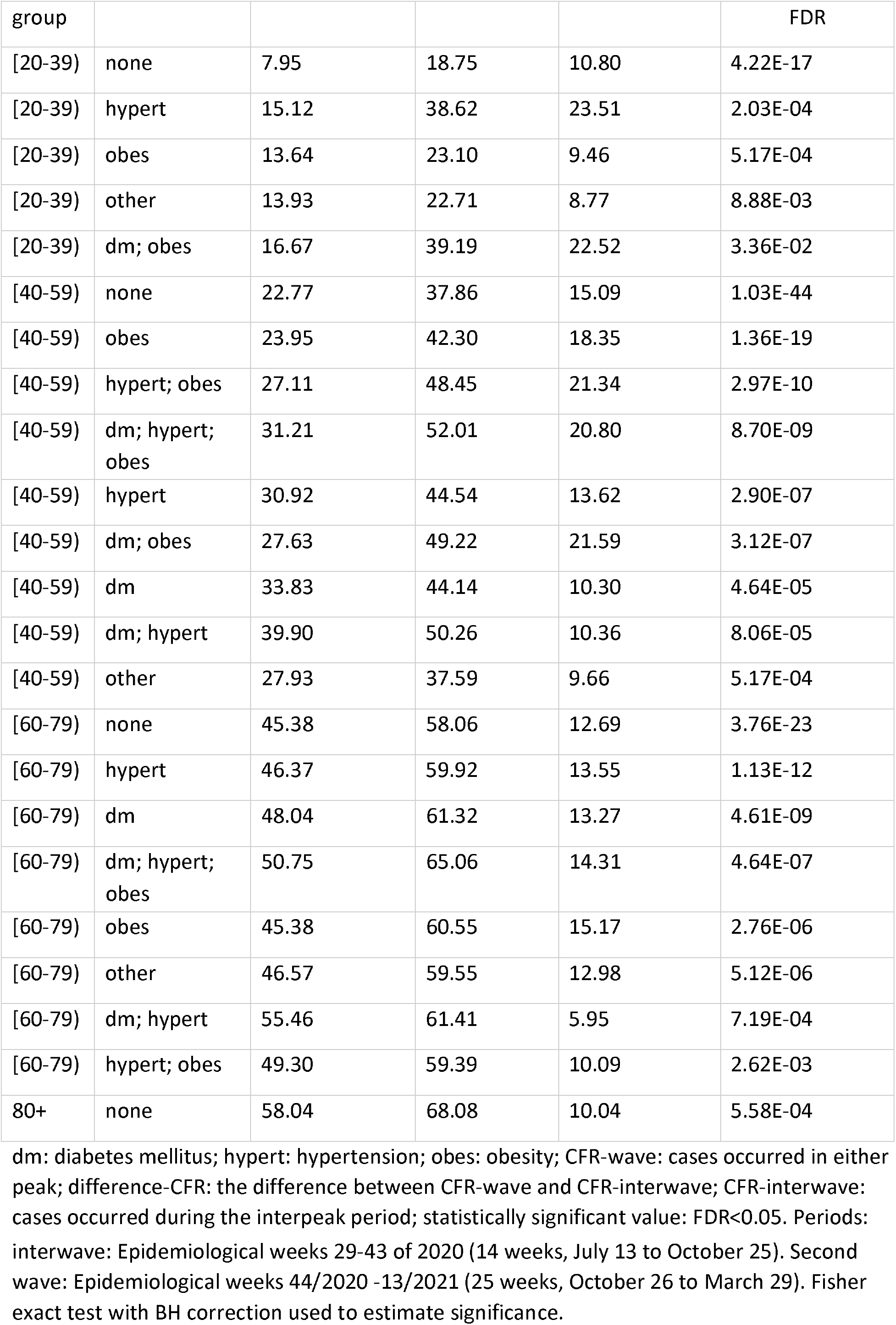
Lethality comparison between the interwave period and the second wave among hospitalized patients with COVID-19.

### Correlation of hospital occupancy with hospital mortality

Considering that lethality increases during periods of higher transmission and demand for hospitalization, we calculated the Pearson correlation between weekly CFR for hospitalized patients and hospital occupancy. Figure 2 shows the time series for CFR and hospital occupancy. We observed that lethality across all age groups decreased as hospital occupation decreased at the end of the first wave, but then increased again during the second wave. Table 10 shows the correlation between weekly hospital occupation and CFR per age group, the correlation values were high in all age groups but the highest was seen in the 60 to 79 age group.

**Table 10:**
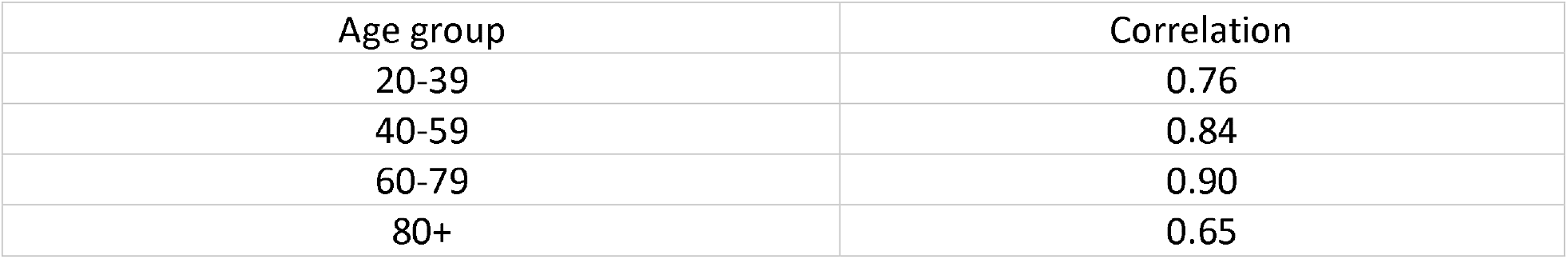
Correlation between weekly hospital occupation and case fatality rate per age group.

**Figure 2.**
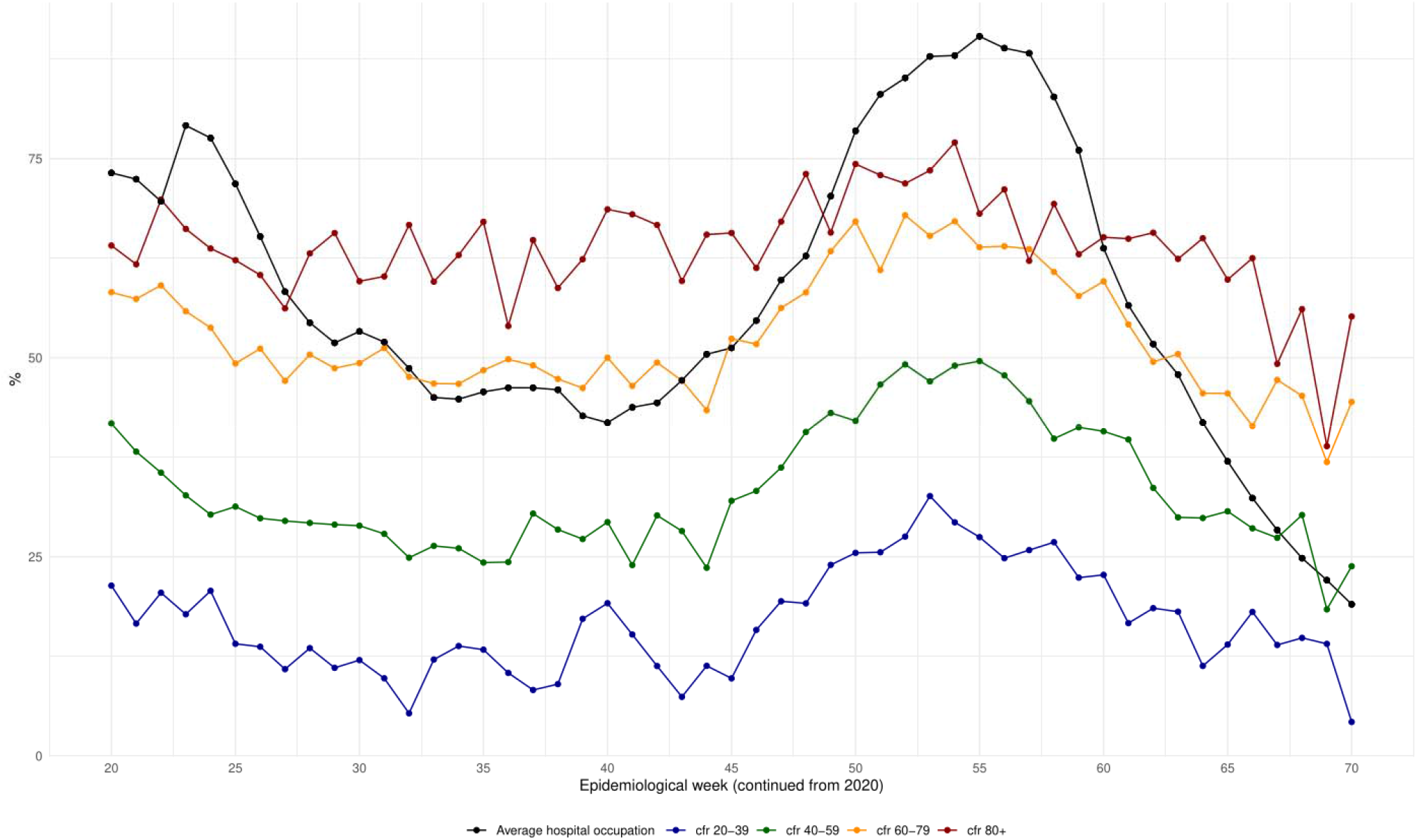
Pearson correlation between weekly case fatality rate (CFR) for hospitalized patients and hospital occupancy. Lethality increases during periods of higher transmission and demand for hospitalization. It was observed that lethality across all age groups decreased as hospital occupation improved at the end of the first wave and increased again during the second wave.

## DISCUSSION

The data for this study come from a population of more than two million people analysed and reveal that even though a considerable increase was observed in the number of cases of infection (+472%), in the number of hospitalized subjects (+85.9%), and the number of hospitalized subjects and deaths (+102.4%) in all age groups between both waves, the probability of death diminished in the general population during the period studied, which is most probably due to the greater number of subjects studied after the first wave. Nevertheless, when analysing only those hospitalized individuals, with or without comorbidities, the CFR was high (50.2%). The probability of death increased considerably in all age groups, and this increase was more noticeable in individuals with previously identified comorbidities (DM, hypertension, or obesity). After concluding this analysis, we were struck by the increased probability of death in the groups of individuals without comorbidities, particularly in the groups aged 20 to 39 years and 40 to 59 years during wave episodes. This indicated to us that the youngest individuals had a very significant risk of death due to the severity of the disease which had led to their hospitalization. This result forces us to maintain the protective measures to avoid infection by the SARS-Cov-2 virus in the entire population, to intensify the vaccination program in young adults and adolescents and improve the medical care of those individuals requiring hospitalization, but with emphasis on the groups that we have identified as having significant risk.

The German team, Staerk et al. [21] described how effective IFRs (infection fatality rates) were estimated to vary over time as the age distributions of confirmed cases and estimated infections were changing during the pandemic. Alimohamadi et al. reported an interesting global analysis [22] of the CFR of COVID-19: the overall pooled CFR of COVID-19 was 10.0% (95% CI: 8.0-11.0). The pooled CFR of COVID-19 among the general population was 1.0% (95% CI: 1.0-3.0); while in hospitalized patients it was 13.0% (95% CI: 9.0-17.0). The pooled CFR in patients admitted into intensive care units was 37.0% (95% CI: 24.0-51.0), and in patients aged over 50, it was 19.0% (95% CI: 13.0-24.0). In our study, we observed a very high CFR in hospitalized COVID-19 patients (50.2%) because of the severity of the infection and its complications, although the high CFR can also be explained by the harmful effect of the saturation of hospital services occurring during the two waves (as can be seen in Figure 2) affecting people under 80 years of age, especially those between 20 and 59 years of age (as shown in Table 9). Our group has previously reported that 45% of patients who did not survive in a tertiary care hospital were not able to be admitted to the ICU due to the lack of availability of ICU beds.[23] Therefore, the mortality rate over time was mainly due to the availability of ICU beds, indirectly suggesting that overcrowding was one of the main contributing factors to hospital mortality. Above all, in this study, the risk observed in people of these age groups (under 60 years) without associated comorbidities is striking.

The risk of SARS-COV-2 infection associated with being over 60 was reported early in the course of the pandemic [24]. Additionally, in both China and the USA, DM, arterial hypertension, and obesity were reported as comorbidities that increase the risk of severe disease, hospitalization, and death.[25] Also early on, a respiratory-onset viral infection was identified as generating progressive multi-organ disease leading initially to respiratory failure and later to multi-organ failure.[26] All of the above are related to an inflammatory phenomenon called a cytokine storm [27], which causes extensive tissue damage with acute functional deterioration and specific organic sequelae in certain groups of patients.[28]

Early on, in Mexico and Latin America, several groups highlighted the rapid progression of patients with COVID-19 [12, 29-32] in coexistence with comorbidities such as DM, high blood pressure, and obesity, as being a worrying situation due to the high prevalence of overweight/obesity, metabolic syndrome and DM in the Mexican population as per information collected by the latest ENSANUT.[8] Then again, we have observed that a good proportion of COVID-19 patients are people in their fourth and fifth decades of life, younger than the patients in the USA reports. Notably, we observed an 85.9% increase in the number of hospitalizations between the two outbreaks, indicative of the severity of the pandemic in the CDMX and evidence of the intense transmission that occurred in that period, there being a notable predominance of a variant designated B.1.1.519. We observed an overlap of the second wave (October 2020 to March 2021) with the emergence of variant B.1.1.519, which represented 74.3% of the sequences generated in CDMX (2296/3092) from November 2020 to May 2021, distributed evenly across CDMX.[15]

We observed that across all age groups, there was an increase in hospitalizations during the second wave, compared with the first wave. The relative growth in the number of hospitalizations as a function of age ranged from a +36.2% increase (from 3261 to 4442) in the 20-39 age group up to a +171.37% increase (from 1544 to 4190) in the 80+ group, as shown in Figure 1. This is evidence that all age groups contributed to the greater pressure on hospital occupation during the second wave, with the most notably increasing demand being in the older-aged groups. Similarly, we observed an increase in the number of deaths and an increase in the probability of death in all age groups with or without comorbidities as described in Table 4. We observed that the probability of dying among individuals with DM, obesity, or arterial hypertension and COVID-19 in the general population was initially reduced in all age groups because of the +800% increase in the number of subjects tested during the study when comparing first and second waves. However, the probability of death in individuals with DM, hypertension, or obesity who were hospitalized increased during the second wave in all age groups, confirming the associated risk observed among the population.

As a result of this careful analysis, we have been able to identify the increased risk of complications and death, between the first and second waves, in all groups of hospitalized patients without comorbidities, from 3.74% in the group aged 20 to 40 to 9.34% in the group of patients aged 40 to 60, which reveals the risk to this population, primarily considered to be healthy. In the healthy population, the appearance of the multisystem inflammatory syndrome has been widely described, especially in children,[33] although at the beginning of the pandemic, it was confused with Kawasaki syndrome. A similar pattern has been described in adults from various regions of the world, in whom a picture of shock rather than respiratory failure [33,34] has been observed, associated with a high risk of death. In such patients, the PCR tests may be negative because it can manifest during the acute phase of the disease or as a postinfectious condition. This clinical scenario has been observed in Mexican children and reported by the group of the National Institute of Paediatrics. [35] In this report, 62% of the studied population did not report previous diseases and it is conceivable that a fraction of this population had presented with this multisystem inflammatory syndrome, however, we recognize that we cannot affirm that this would have been the diagnosis. Therefore, more research is required to ascertain if this condition exists in the Mexican population.

Attention has recently been drawn to the appearance of severe COVID-19 in groups of people without comorbidities. In fact, the study recently published by ISARIC WHO Clinical Characterization Protocol (UK) draws attention to the appearance of serious disease in this group of people and their need for hospitalization. In consequence, they have pointed out that it is helpful to look for other outcomes such as disability or chronic damage that may appear in the members of this group who survive but may develop complications in the medium and long term.[7] Recently, loci and genes associated with susceptibility to COVID-19 or to severe COVID-19 have been identified through the use of comprehensive GWAS (Genome-wide association studies) or analysis of genome, exome, or candidate genes.[36,37] Some of the loci reported include ABO blood groups, [36] ACE2 receptor,[38] TMPRSS2 (host transmembrane serine protease), [38] various HLA, [39] APOE,[40] and IFITM3 (interferon-induced transmembrane protein-3) [41] alleles. On the other hand, rare variants in genes encoding members of the type I/III interferon (IFN) pathway showed a higher estimated risk of severe COVID-19,[42] and some rare and deleterious germline variants in the X chromosome Toll-like receptor 7 (TLR7) gene have also been reported in young, healthy men with severe COVID-19,[43], these being possible immune defects that may explain the COVID-19 severity among these otherwise healthy people.

Based on our results, the results of the study carried out by our group in the tertiary care hospital [23] and the results of other recently published studies,[30] we consider that it is necessary to carry out the following measures to avoid an increase in the rates of lethality in hospitalized patients. The first measure is, without a doubt, to intensify public health measures shown to decrease transmission (use of face masks, social distancing, and continuous application of hygiene measures). The second is the intensification of vaccination among different age groups progressively until the entire population is covered. In this way, we can significantly reduce the number of cases of hospitalization, episodes of aggravation of disease, transfers to intensive care units, and the need for endotracheal intubation, as well as avoid cases of disability and death. (The data for Mexico City are currently under analysis). The third measure is the promotion of early care to recognize patients with risk comorbidities or those individuals with clinical manifestations or alterations in laboratory tests, indicative of severity or progressive deterioration.[44]

As regards the limitations and strengths of this study, its main limitation was that the recruitment method of the persons infected by SARS-CoV-2 was modified from symptomatic during the first wave to both asymptomatic and symptomatic in the other two periods, in addition to the modification of the diagnostic procedure from tPCR to antigen detection for the screening method. This condition prevented an adequate calculation of the infection fatality rate since the inclusion of symptomatic individuals predominated in the screening system. Another significant limitation was being unable to follow up on all the contacts of at least hospitalized patients to gain a better understanding of the severity of transmission. Furthermore, there were problems registering deaths and difficulties completing the registration, especially during the first wave. Therefore, for patients who died without a PCR-SARS-CoV-2 test, a ‘probable COVID-19’ code was added to include those patients without a confirmatory test. On the other hand, although this was not designed as a population-based study, the study’s main strength remains in its inclusion and consideration of all hospitalized patients, exploring their outcomes and carrying out the entire probabilistic and Bayesian analysis. Although the SISVER database might include some ‘measurement uncertainty’, the Bayesian model is not affected by this kind of ‘uncertainty’ since the potential biases were incorporated into the priors from the start. Thus, the resulting analysis model was in some sense ‘strengthened’ by this strategy.

## CONCLUSION

This observational study revealed a considerable increase in the number of cases (+472%) of infection plus an +85.9% increase in hospitalized cases in all age groups between the first and second waves. Additionally, 12.8% of the infected persons were hospitalized because of severe COVID-19, with a high mortality rate among the hospitalized patients (>50%). Case Fatality Rates among hospitalized patients were higher during the peaks than during the interwave period. A higher probability of death was observed among hospitalized patients who suffered from diabetes, obesity, or hypertension. This probability increased during periods of higher hospital demand. It should be noted that this increase in the risk of death was observed in all age groups, regardless of the presence or absence of comorbidities.

## Data Availability

All data produced in the present study are available upon reasonable request to the authors

https://www.sinave.gob.mx/

https://sisver.sinave.gob.mx/influenza/

## SUPPLEMENTARY MATERIAL

**Table S1.**
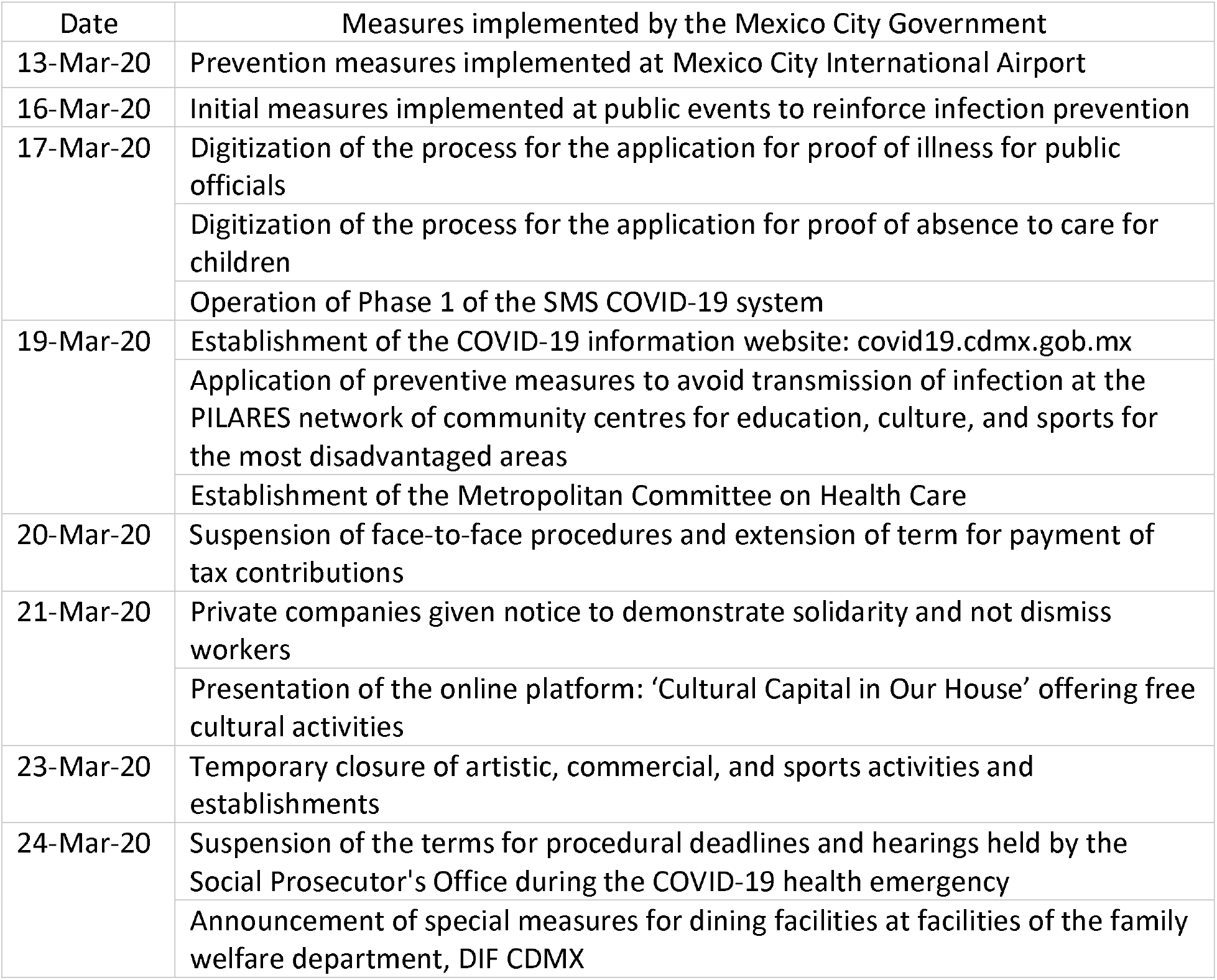
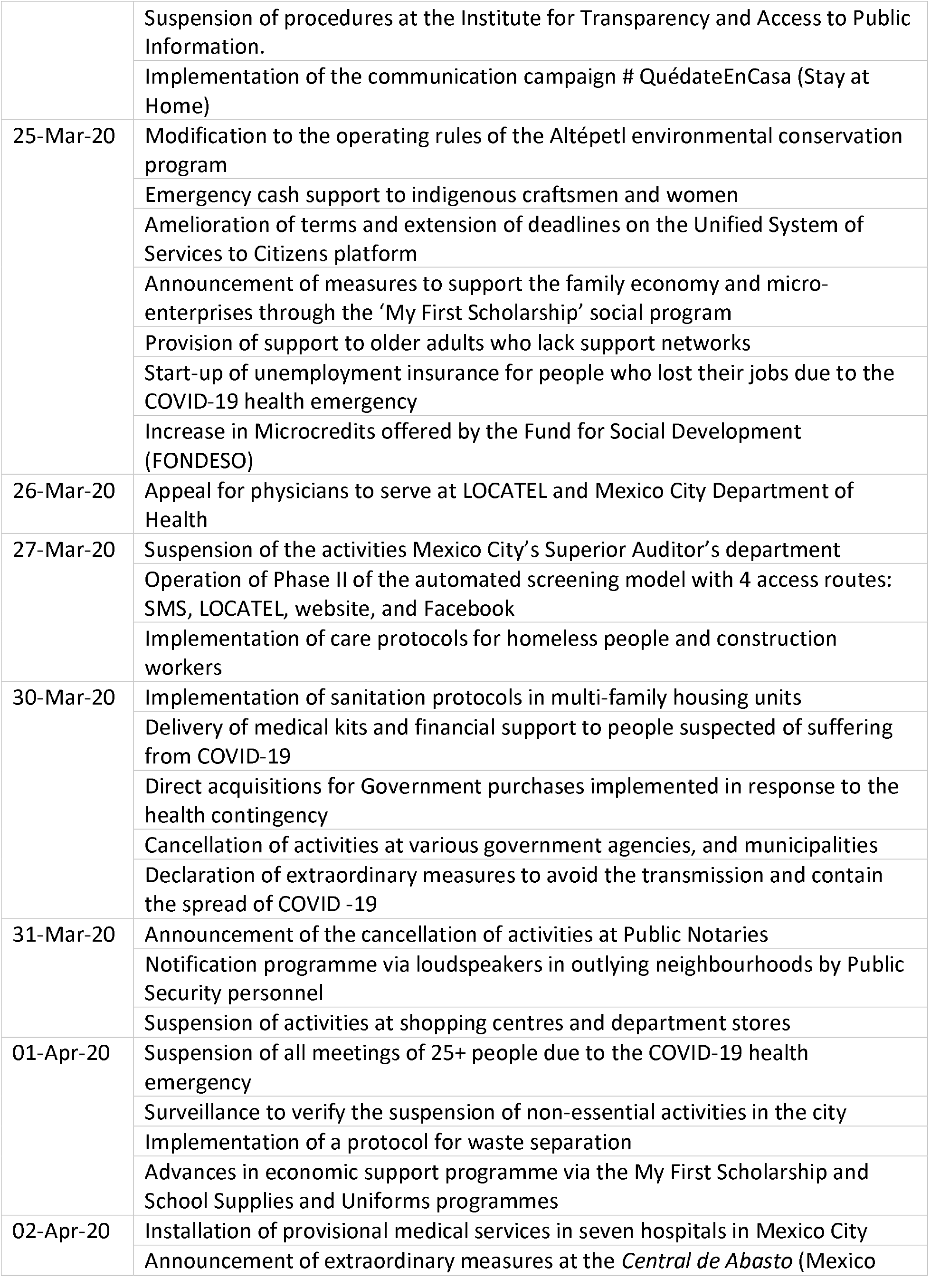
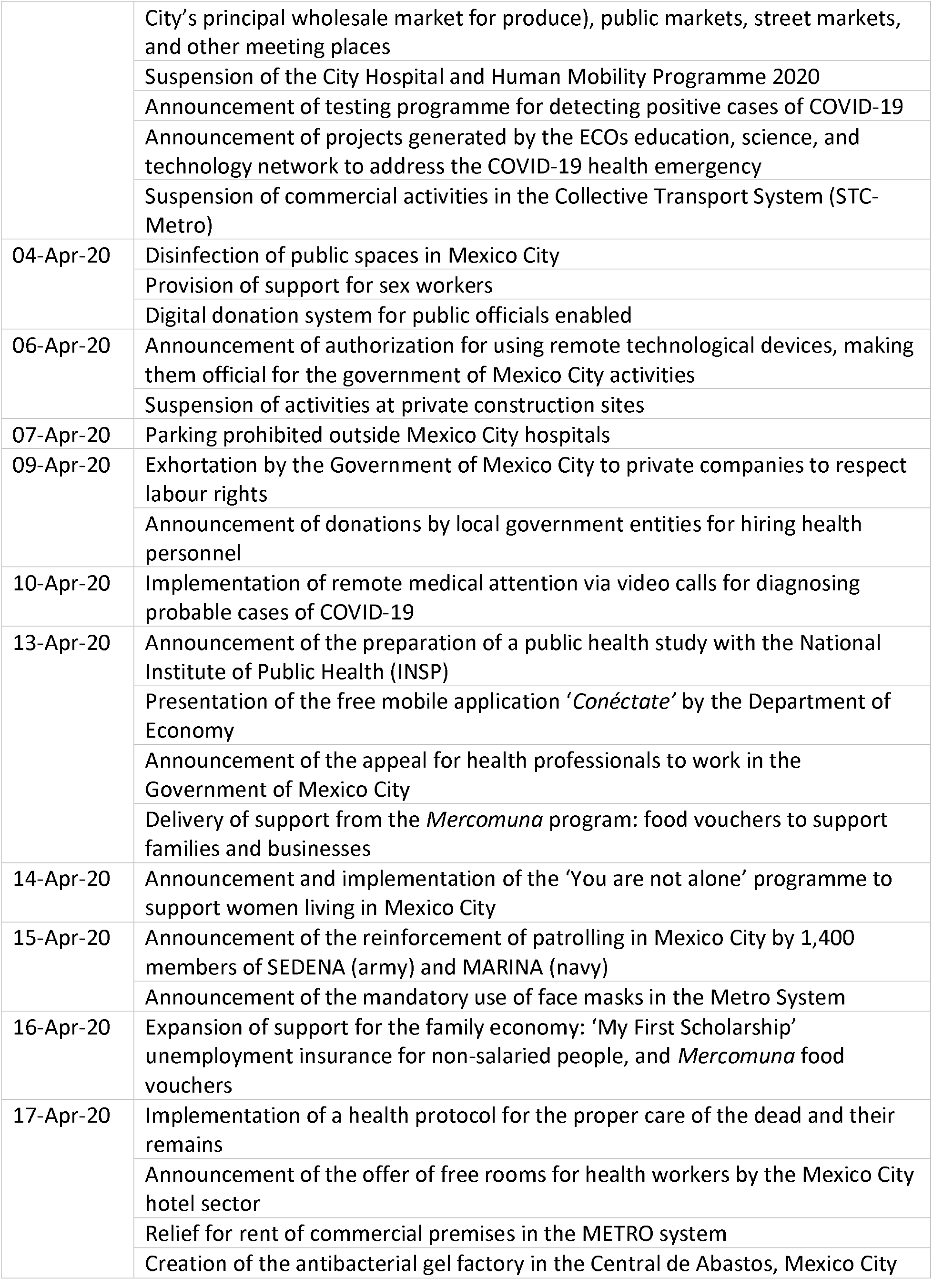
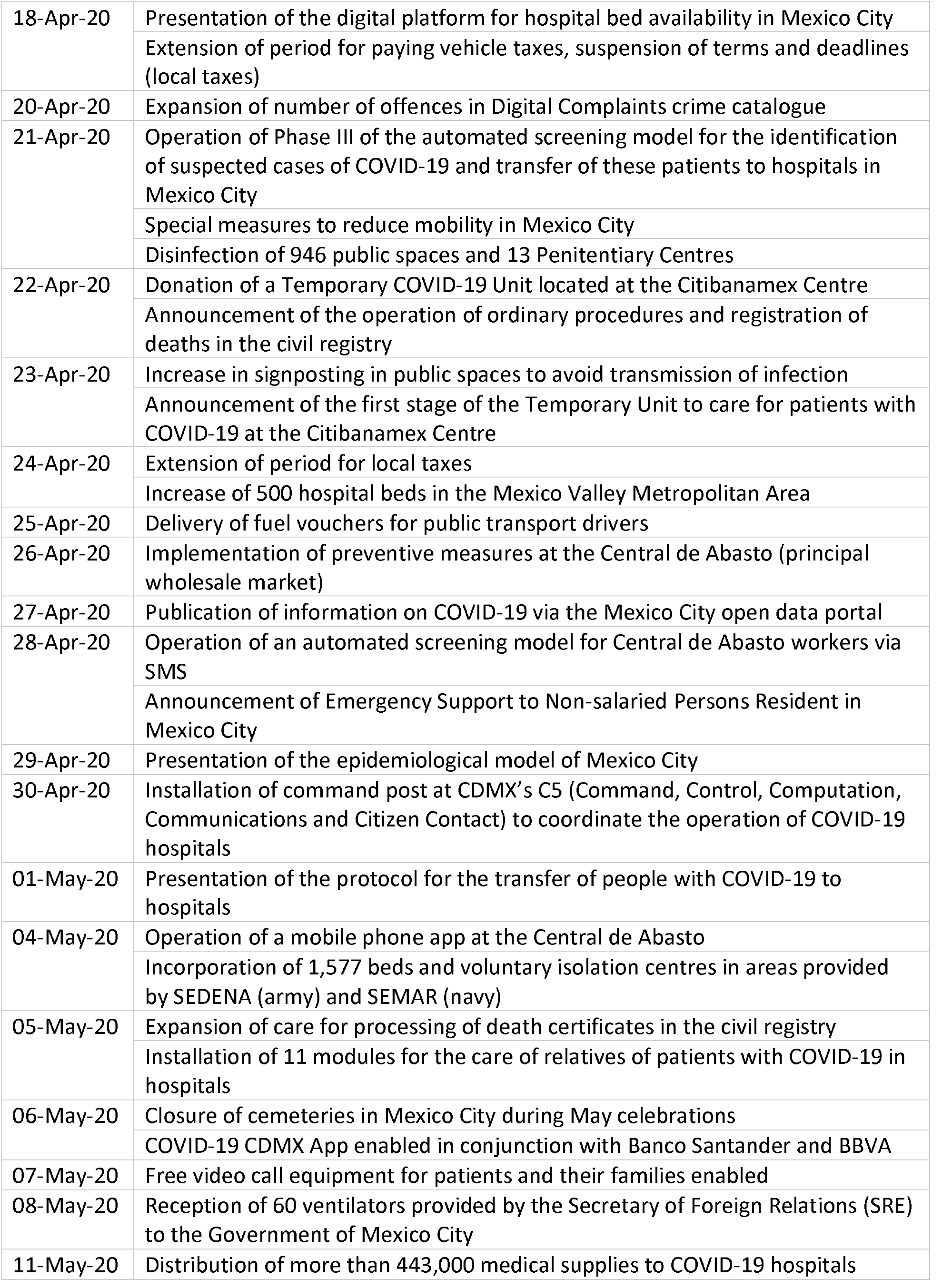
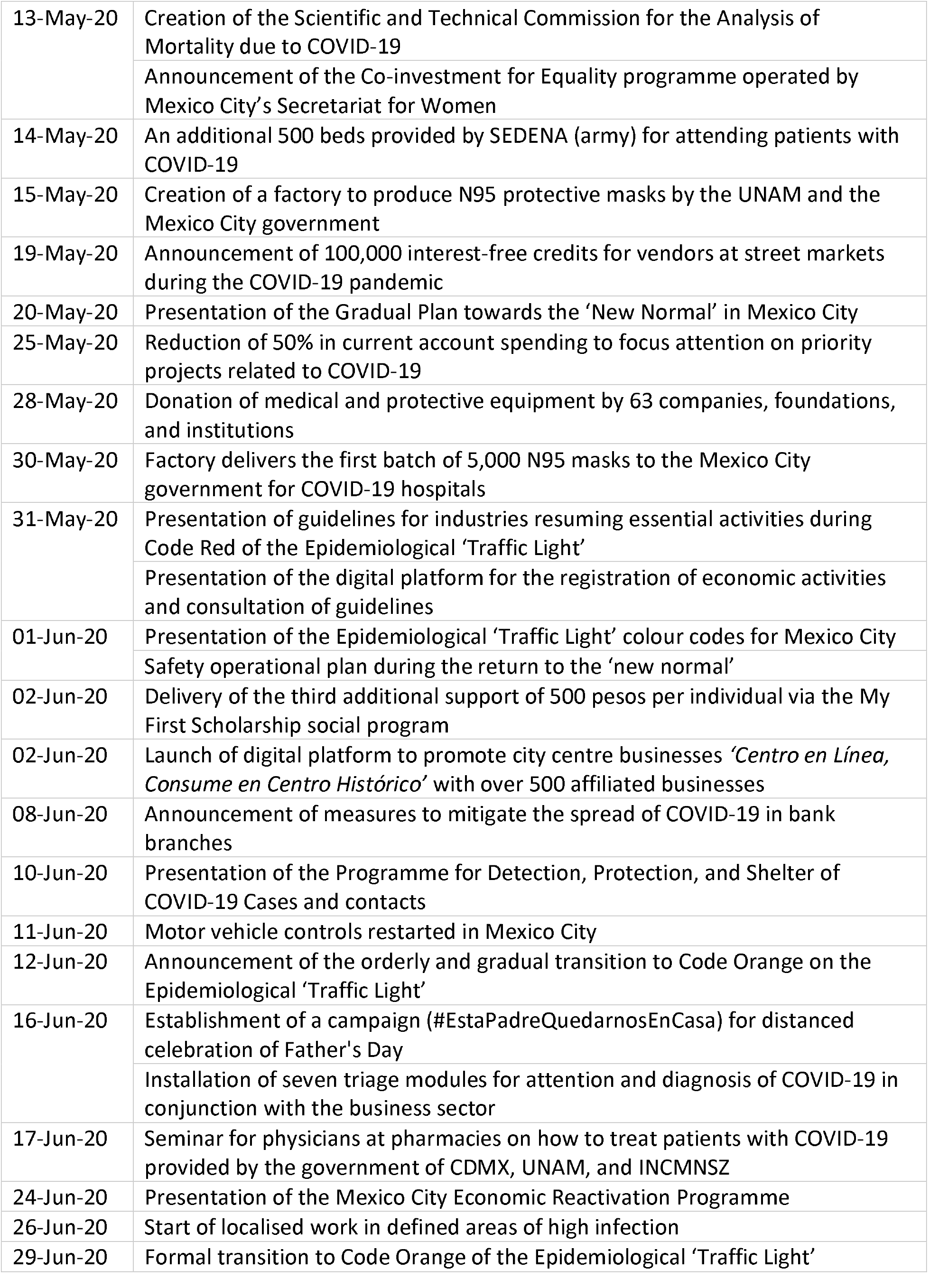
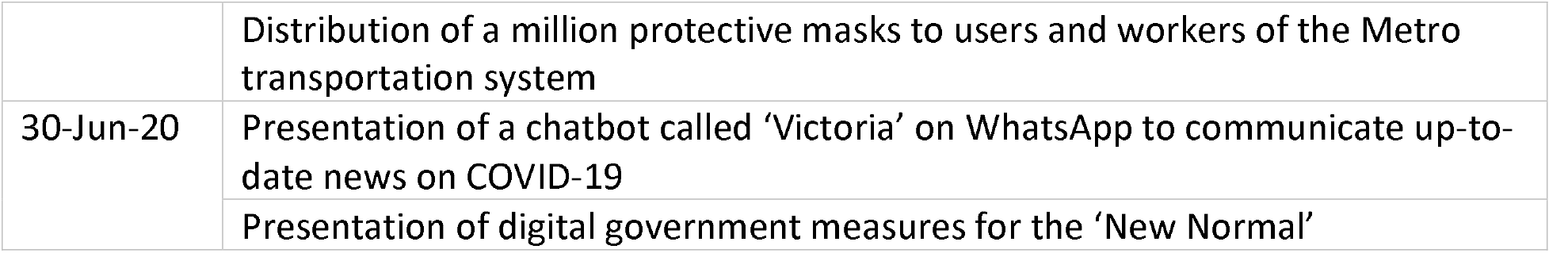
Measures taken to combat the COVID-19 pandemic in Mexico City during the first wave (15 weeks, epidemiological weeks 13 to 28) March 23 to July 6, 2020.

**Table S2:** Measures taken to combat the COVID-19 pandemic in Mexico City during the interwave period (14 weeks, from epidemiological weeks 29 to 43) July 13 to October 19, 2020.

| Date | Measures implemented by the Government of Mexico City |
| --- | --- |
| 06-Jul-20 | Presentation of additional measures for department stores and shopping centres |
| 10-Jul-20 | Presentation of the Responsible and Protected Homes Programme as well as measures implemented in neighbourhoods to be classified as Code Red, having larger numbers of COVID-19 cases |
|  | Strengthening of procedures for the delivery of kits under the Responsible and Protected Household Program |
| 13-Jul-20 | Presentation of the first 34 health kiosks providing medical advice |
| 14-Jul-20 | Registration of beneficiaries initiated for the 'My First Scholarship' programme using the electronic application |
| 22-Jul-20 | Announcement of attention in 36 priority neighbourhoods due to COVID-19 infections |
| 29-Jul-20 | Application of diagnostic tests mandated for 3% of employees in establishments with a staff of 100 or more workers |
| 31-Jul-20 | Announcement that the provision of 'Support for School Supplies and Uniforms' will be carried out in two stages |
| 07-Aug-20 | Presentation of the ReABRE Program: Reopening of bars and restaurants |
| 12-Aug-20 | Delivery of the *Leona Vicario* scholarship to children and adolescents orphaned by COVID-19 |
| 11-Sep-20 | Announcement that tests results may be viewed using the CDMX app, speeding up notifications |
| 15-Oct-20 | Measures to strengthen detection, protection, and shelter for COVID-19 cases |

**Table S3.**
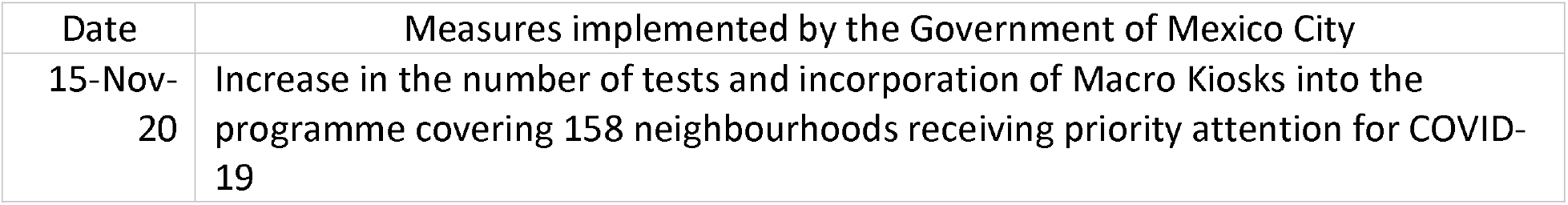
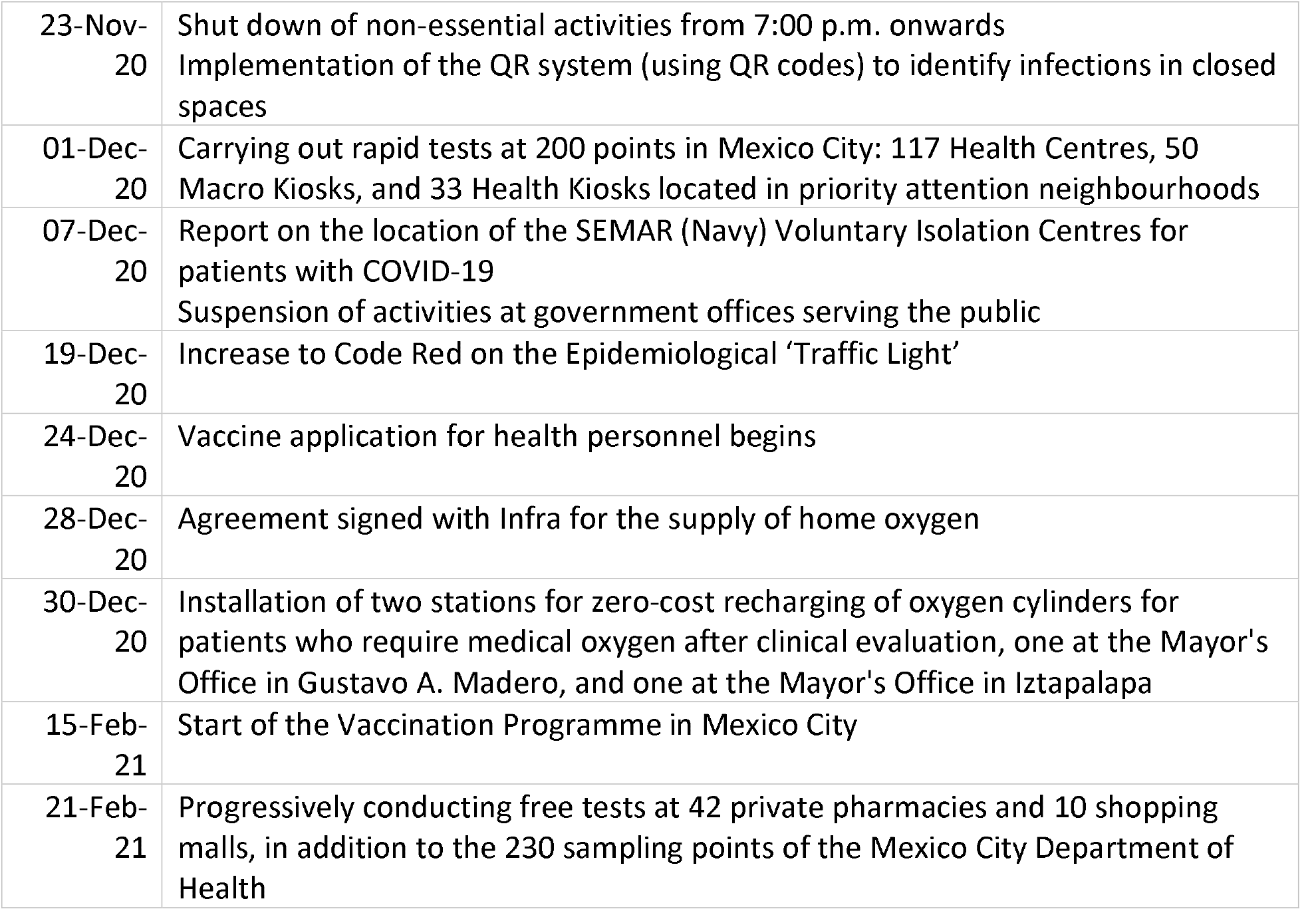
Measures implemented to combat the COVID-19 pandemic in Mexico City during the second wave (25 weeks, between October 26, 2020, to March 29, 2021).

**Table S4.** Comparison of lethality (in ambulatory and hospitalized patients) between interwave period and the second wave.

| Age group (years) | Comorbidities | CFR_interwave | CFR_2nd wave | Difference-CFR | FDR |
| --- | --- | --- | --- | --- | --- |
| [60,79) | dm; hypert | 27.72 | 22.06 | -5.66 | 9.72E-08 |
| [60,79) | none | 15.25 | 12.78 | -2.46 | 2.24E-06 |
| [40,59) | obes | 4.48 | 6.10 | 1.62 | 2.26E-04 |
| [40,59) | hypert; obes | 6.43 | 9.82 | 3.40 | 7.25E-04 |
| [40,59) | dm; hypert; obes | 11.45 | 16.49 | 5.03 | 3.53E-03 |
| [80,100) | dm; hypert | 44.61 | 36.19 | -8.41 | 8.85E-03 |
| [60,79) | obes | 18.61 | 23.20 | 4.58 | 2.20E-02 |
| [80,100) | none | 34.56 | 29.18 | -5.38 | 3.09E-02 |
| [40,59) | dm; hypert | 13.63 | 11.28 | -2.35 | 3.38E-02 |
| [60,79) | dm | 20.11 | 17.48 | -2.62 | 4.13E-02 |

dm: diabetes mellitus; hypert: hypertension arterial; obes: obesity; CFR-wave: cases occurred in either peak; difference-CFR: the difference between CFR-wave and CFR-interwave; CFR-interwave: cases occurred during the interpeak period; statistical significant value: FDR<0.05. Periods: interwave: Epidemiological weeks 29-43 of 2020 (14 weeks, July 13 to October 25). Second wave: Epidemiological weeks 44/2020 -13/2021 (25 weeks, October 26 to March 29). Fisher exact test with BH correction used to estimate significance. Positive values in Difference-CFR represent **increases** in lethality. Negative values in Difference-CFR represent **decreases** in lethality.

**Table S5.** Frequencies of people with diabetes mellitus, positive for SARS-CoV-2 infection, hospitalized and deceased during the first wave (15 weeks, between March 23 to July 12, 2020).

| Age group (years) | People with diabetes | People with SARS-CoV-2 infection | People with COVID-19 and hospitalized | Deceased people with COVID-19 | People with COVID-19, hospitalized and deceased |
| --- | --- | --- | --- | --- | --- |
| 20-39 | 1970 | 940 | 314 | 123 | 106 |
| 40-59 | 11812 | 6017 | 2754 | 1399 | 1245 |
| 60-79 | 9225 | 5299 | 3321 | 2202 | 1980 |
| >80 | 1316 | 703 | 490 | 374 | 332 |

**Table S6.** Frequencies of people with diabetes mellitus, positive for SARS-CoV-2 infection, hospitalized and deceased during the interwave period (14 weeks, July 13 to October 25, 2020).

| Age group (years) | People with diabetes | People with SARS-CoV-2 infection | People with COVID-19 and hospitalized | Deceased people with COVID-19 | People with COVID-19, hospitalized and deceased |
| --- | --- | --- | --- | --- | --- |
| 20-39 | 2661 | 1001 | 166 | 37 | 35 |
| 40-59 | 16669 | 6607 | 1723 | 643 | 598 |
| 60-79 | 13409 | 5790 | 2620 | 1473 | 1373 |
| >80 | 1615 | 751 | 475 | 328 | 304 |

**Table S7.** Frequencies of people with diabetes mellitus, positive for SARS-CoV-infection, hospitalized and deceased during the second wave (25 weeks, between 26 October 2020 to 29 March 2021).

| Age group (years) | People with diabetes | People with SARS-CoV-2 infection | People with COVID-19 and hospitalized | Deceased people with COVID-19 | People with COVID-19, hospitalized and deceased |
| --- | --- | --- | --- | --- | --- |
| 20-40 | 10850 | 3407 | 374 | 145 | 137 |
| 40-60 | 67455 | 22818 | 3993 | 2079 | 1920 |
| 60-80 | 52223 | 20483 | 6750 | 4513 | 4178 |
| >80 | 5442 | 2458 | 1226 | 903 | 839 |

**Table S8.** Frequencies of people with hypertension, positive for SARS-CoV-2 infection, hospitalized and deceased during the first wave (15 weeks, March 23 to July 12, 2020).

| Age group (years) | People with hypertension | People with SARS-CoV-2 infection | People with COVID-19 and hospitalized | Deceased people with COVID-19 | People with COVID-19, hospitalized and deceased |
| --- | --- | --- | --- | --- | --- |
| 20-40 | 2691 | 1124 | 297 | 115 | 94 |
| 40-60 | 13836 | 6548 | 2627 | 1269 | 1125 |
| 60-80 | 11714 | 6548 | 3833 | 2515 | 2248 |
| >80 | 2293 | 1218 | 817 | 623 | 551 |

**Table S9.** Frequencies of people with hypertension, positive for SARS-CoV-2 infection, hospitalized and deceased during the interwave period (14 weeks, July 13 to October 25, 2020).

| Age group (years) | People with hypertension | People with SARS-CoV-2 infection | People with COVID-19 and hospitalized | Deceased people with COVID-19 | People with COVID-19, hospitalized and deceased |
| --- | --- | --- | --- | --- | --- |
| 20-40 | 3889 | 1313 | 203 | 50 | 46 |
| 40-60 | 21018 | 7772 | 1750 | 627 | 584 |
| 60-80 | 18327 | 7458 | 3074 | 1696 | 1576 |
| >80 | 2913 | 1316 | 764 | 543 | 504 |

**Table S10.** Frequencies of people with hypertension, positive for SARS-CoV-2 infection, hospitalized and deceased during the second wave (25 weeks, between Oct 26, 2020, to March 29, 2021).

| Age group (years) | People with hypertension | People with SARS-CoV-2 infection | People with COVID-19 and hospitalized | Deceased people with COVID-19 | People with COVID-19, hospitalized and deceased |
| --- | --- | --- | --- | --- | --- |
| 20-40 | 16464 | 4815 | 425 | 181 | 171 |
| 40-60 | 86466 | 27486 | 4141 | 2185 | 1999 |
| 60-80 | 72959 | 27040 | 8257 | 5446 | 5049 |
| >80 | 10082 | 4392 | 2091 | 1547 | 1437 |

**Table S11.** Frequencies of people with obesity, positive for SARS-CoV-2 infection, hospitalized and deceased during the first wave (15 weeks, March 23 to July 12, 2020).

| Age group | People with obesity | People with SARS-CoV-2 | People with COVID-19 | Deceased people with | People with COVID-19, |
| --- | --- | --- | --- | --- | --- |

| (years) |  | infection | and<br>hospitalized | COVID-19 | hospitalized<br>and<br>deceased |
| --- | --- | --- | --- | --- | --- |
| 20-40 | 11062 | 4698 | 773 | 230 | 195 |
| 40-60 | 16118 | 7834 | 2600 | 1170 | 1007 |
| 60-80 | 5049 | 2984 | 1673 | 1094 | 958 |
| >80 | 448 | 247 | 152 | 122 | 104 |

**Table S12.**
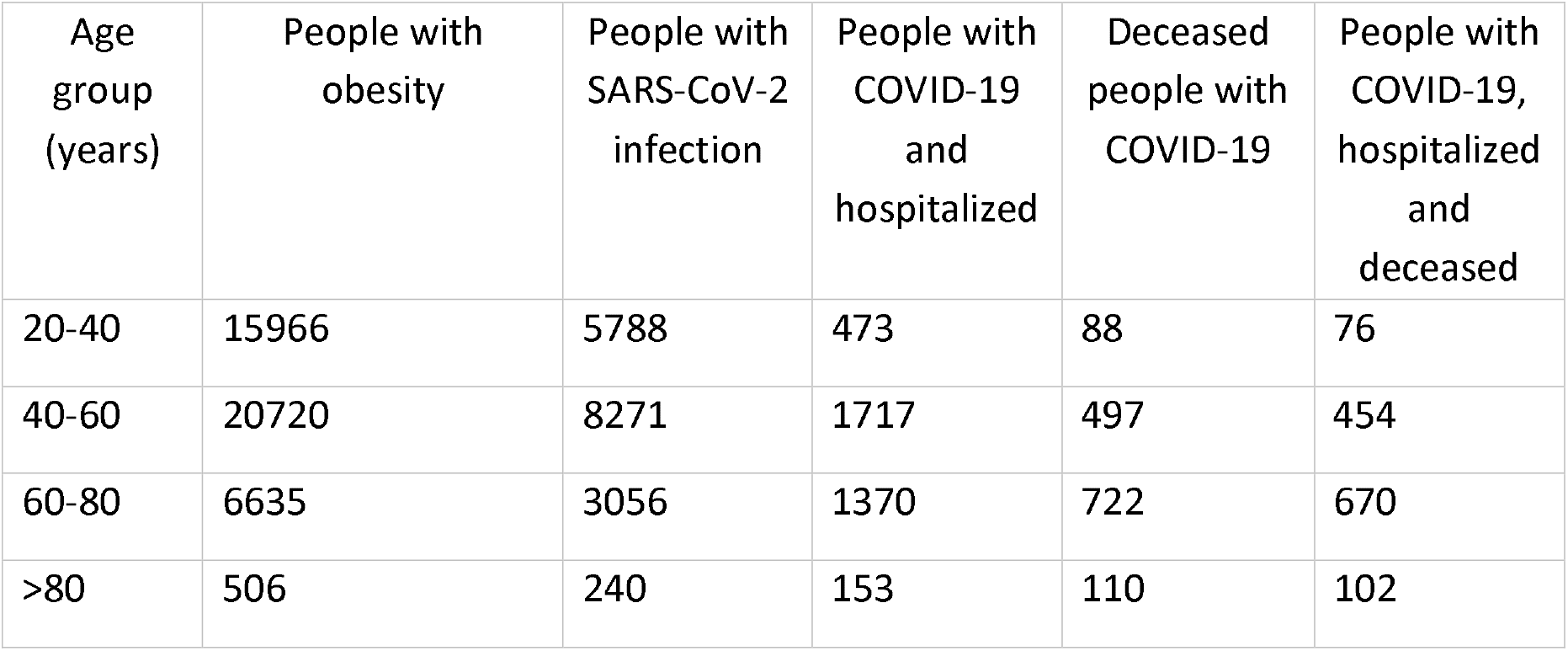
Frequencies of people with obesity, positive for SARS-CoV-2 infection, hospitalized and deceased during the interwave period (15 weeks, July 13 to October 25, 2020).

| Age<br>group<br>(years) | People with<br>obesity | People with<br>SARS-CoV-2<br>infection | People with<br>COVID-19<br>and<br>hospitalized | Deceased<br>people with<br>COVID-19 | People with<br>COVID-19,<br>hospitalized<br>and<br>deceased |
| --- | --- | --- | --- | --- | --- |
| 20-40 | 15966 | 5788 | 473 | 88 | 76 |
| 40-60 | 20720 | 8271 | 1717 | 497 | 454 |
| 60-80 | 6635 | 3056 | 1370 | 722 | 670 |
| >80 | 506 | 240 | 153 | 110 | 102 |

**Table S13.** Frequencies of people with obesity, positive for SARS-CoV-2 infection, hospitalized and deceased during the second wave (25 weeks, between October 26 to March 29, 2021).

| Age<br>group<br>(years) | People<br>with<br>obesity | People with<br>SARS-CoV-2<br>infection | People with<br>COVID-19 and<br>hospitalized | Deceased<br>people with<br>COVID-19 | People with<br>COVID-19,<br>hospitalized and<br>deceased |
| --- | --- | --- | --- | --- | --- |
| 20-40 | 55280 | 16457 | 1004 | 299 | 273 |
| 40-60 | 65931 | 22055 | 3713 | 1851 | 1714 |
| 60-80 | 19573 | 8458 | 3436 | 2258 | 2120 |
| >80 | 1481 | 735 | 436 | 321 | 300 |

**Table S14.**
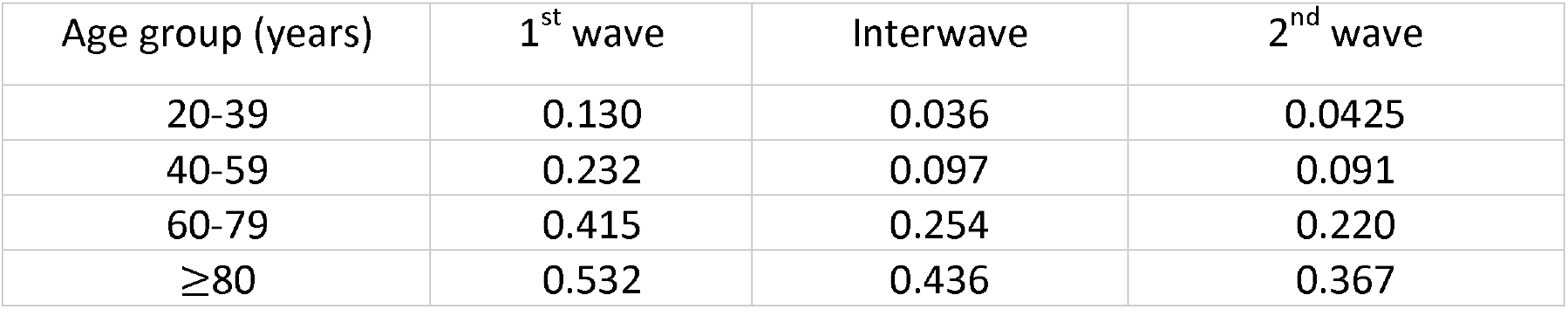
Probability of death of COVID-19 patients with diabetes mellitus according to age groups and pandemic waves.

| Age group (years) | 1<sup>st</sup> wave | Interwave | 2<sup>nd</sup> wave |
| --- | --- | --- | --- |
| 20-39 | 0.130 | 0.036 | 0.0425 |
| 40-59 | 0.232 | 0.097 | 0.091 |
| 60-79 | 0.415 | 0.254 | 0.220 |
| ≥80 | 0.532 | 0.436 | 0.367 |

**Table S15.** Probability of death of COVID-19 patients with hypertension according to age groups and pandemic waves.

| Age group (years) | 1<sup>st</sup> wave | Interwave | 2<sup>nd</sup> wave |
| --- | --- | --- | --- |
| 20-39 | 0.102 | 0.038 | 0.037 |
| 40-59 | 0.193 | 0.080 | 0.079 |
| 60-79 | 0.384 | 0.227 | 0.201 |
| ≥80 | 0.511 | 0.412 | 0.352 |

**Table S16.**
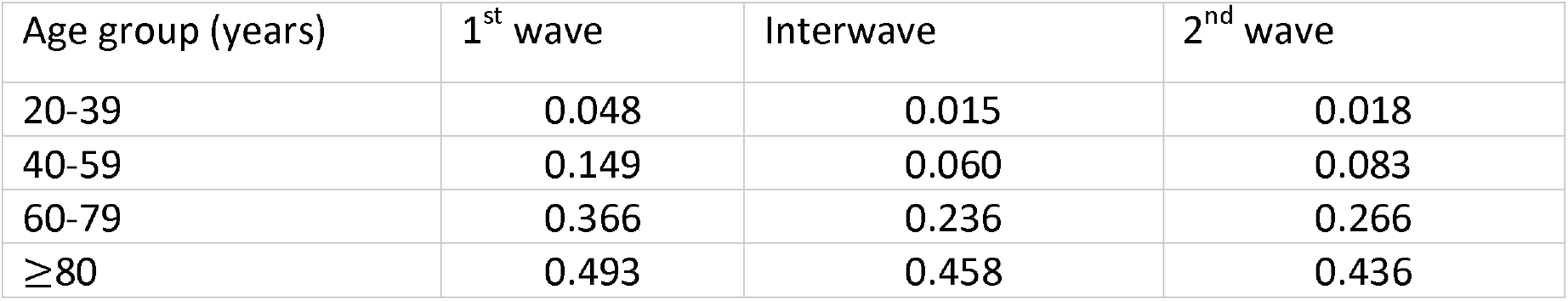
Probability of death of COVID-19 patients with obesity according to age groups and pandemic waves.

| Age group (years) | 1<sup>st</sup> wave | Interwave | 2<sup>nd</sup> wave |
| --- | --- | --- | --- |
| 20-39 | 0.048 | 0.015 | 0.018 |
| 40-59 | 0.149 | 0.060 | 0.083 |
| 60-79 | 0.366 | 0.236 | 0.266 |
| ≥80 | 0.493 | 0.458 | 0.436 |

